# Genomic characterization of *Klebsiella pneumoniae* causing invasive disease in South African infants: observational studies between 2018 to 2023

**DOI:** 10.1101/2025.06.04.25328999

**Authors:** Courtney P. Olwagen, Alane Izu, Shama Khan, Stephanie Jones, Carmen Briner, Gaurav Kwatra, Lara Van der Merwe, Nicholas J. Dean, Vicky L. Baillie, Sana Mahtab, Kimberleigh Storath, Imaan Dunn, Lubomira Andrew, Urvi Rajyaguru, Firdose L. Nakwa, Sithembiso C. Velaphi, Jeannette Wadula, Renate Strehlau, Anika M. van Niekerk, Niree Naidoo, Yogandree Ramsamy, Mohamed Said, Robert G.K. Donald, Raphael Simon, Ziyaad Dangor, Shabir A. Madhi

## Abstract

**Background:** *Klebsiella pneumoniae* (KPn) is a leading cause of invasive bacterial disease in African children, albeit with a scarcity of genotypic characterization.

**Methods:** Invasive KPn isolates collected from young infants ≤90 days of age through observational surveillance across six hospitals in South Africa between March 4^th^, 2019 to February 27^th^, 2021, and between May 13^th^, 2022 to October 31^st^, 2023 were sequenced. Postmortem Kpn isolates attributed in the causal pathway to death, collected from decedents ≤90 days of age and stillbirths between February 15^th^, 2018 to April 18^th^, 2023, were also sequenced.

**Results:** Three hundred and thirty-seven isolates (226 identified during hospital surveillance and 111 from postmortem sampling) were included in the final analysis. Genomic analysis identified 85 distinct clonotypes. Sequence type (ST) 17 (22.0%,74/337) predominated, followed by ST39 (12.7%,43/337). The dominant K-locus (KL) identified were KL25 (24.0%,81/337), KL2 (14.5%,49/337), KL149 (13/4%,45/337), and KL102 (9.5%,32/337), and the dominant O-antigens detected were O1ab (48.4%,149/310), O5 (19.9%,67/337), and O4 (7.7%,26/337). Eighty-five percent (287/337) of the KPn isolates harboured multi-drug resistant (MDR) genes, including 32.9% (111/337) to carbapenems. The oxacillinase-type β-lactamase-48 (bla_OXA-181_) gene was detected in 26.4% (89/337) of the isolates, while the New Delhi metallo β-lactamase-five (bla_NDM-5_) and -one (bla_NDM-1_) detected in 2.1% (7/337) and 0.3% (1/337) of isolates, respectively.

**Conclusions:** Although a wide diversity of strains were associated with Kpn invasive disease in South African children, over 80% of the cases were attributed to eleven K loci. The findings from this study could be useful in selecting KPn antigen targets for potential vaccine candidates.

## BACKGROUND

Sepsis is a leading cause of death in young infants (≤90 days of age) in low-middle-income countries (LMIC) [1]. Global estimates indicate that the incidence of sepsis among young infants ranges from 1.3 to 3.9 million cases, with sepsis-related complications leading to approximately 400,000 to 700,000 deaths each year [2], 42% of which occur within the first week of life [3]. The burden of invasive bacterial disease is likely underestimated in LMIC, due to limited access to health facilities and laboratory resources to determine the cause of sepsis [4]. In sub-Saharan Africa, the reported incidence of neonatal sepsis ranges from 4.5 to 21 per 1000 live births, with a case-fatality risk of 27% to 56% [4]. In contrast, the estimated incidence of neonatal sepsis is 0.8 to 1 per 1000 in the United States, where the case-fatality risk is 3% to 19% [5, 6]. Moreover, there are an estimated 2 million annual stillbirths globally, >98% of which occur in LMIC [7].

Despite the prominent role of *Klebsiella pneumoniae* (KPn) in causing infant sepsis and death, most studies from LMICs report on the burden of KPn as part of a group with other Gram-negative bacteria. Nevertheless, *K. pneumoniae* has been reported as a common cause of presumed hospital-acquired infections (pHAI) and outbreaks in sub-Saharan Africa, including South Africa [8–12], and is often associated with resistance to multiple antibiotic classes. Investigation of the causes of childhood deaths using post-mortem minimally invasive tissue sampling (MITs), implicated sepsis in the causal pathway to death in two-thirds of decedents, the major pathogens due to hospital-acquired organisms including KPn [13, 14]. There is a paucity of data on the genetic characterization of KPn isolates from LMIC, which could assist in delineating the epidemiology of illness and identify targets for potential interventions including vaccine development.

This study aimed to genetically characterize KPn isolates associated with invasive disease, including fatal cases, over six years in South African infants.

## METHODS

### Study design and population

The analyzed KPn isolates were collected across four observational studies which included KPn surveillance. The first study was undertaken from March 4^th^, 2019 to February 7^th^, 2021 across six hospitals as described [15]. The second study was conducted from May 22^nd^, 2022 to December 31st, 2023, and included surveillance at the same six sites as the earlier study. In both studies, detailed in the Supplementary text, infants hospitalized for presumed serious bacterial infection were evaluated at the discretion of the attending physician, which included blood culture and a lumbar puncture in young infants with suspected meningitis. Microbiological testing was done at the National Health Laboratory Service (NHLS), the sole laboratory servicing public health facilities in South Africa, which has standardized equipment across its laboratories. As standard of care, the recommended empiric antibiotic treatment for neonates admitted with suspected sepsis is ampicillin and gentamicin if hospitalized within 72 hours of birth or hospital admission. In some instances, repeat blood and CSF cultures were undertaken during hospitalization to evaluate the antibiotic response and/or investigate newly suspected sepsis events. Demographic and risk factors for invasive KPn disease were evaluated at admission, including data on the child’s gender, mode of delivery, and gestational age at delivery.

We also analyzed KPn isolates from decedents in which the organism was attributed in the causal pathway of death. The decedents were enrolled between February 15^th^, 2018 and April 18^th^, 2023 from the Child Health and Mortality Prevention Study (CHAMPS) at the Soweto, South Africa site (www.champshealth.org). The CHAMPS study aims to determine the causes of under-five childhood deaths and stillbirths across seven LMICs, including South Africa. Minimally invasive tissue sampling (MITs) is performed within 24 to 72 hours of death and samples including blood, CSF, and lung tissue are sent for culture. For each case, the cause of death (COD) is determined by a multi-disciplinary panel constituted of local experts including neonatologists, paediatricians, microbiologists, and histopathologists. The determination of the cause of death (DeCoDe) panel evaluated all available ante-mortem and post-mortem data and reported on the COD according to WHO recommended guidelines [16].

Additional KPn isolates from decedents not eligible for enrolment into the CHAMPS study, as they were from outside of the Health Demographic Surveillance Site used in the CHAMPS study, were also enrolled between April 17^th^, 2018 to September 14^th^, 2022. The decedents who were enrolled outside of the CHAMPS program had more targeted investigations focused on identifying infectious causes of death (i.e. MITs-lite). The differences in the procedures used for the CHAMPS and MITs-lite surveillance are summarized in Supplementary Table 1. In both post-mortem studies, clinical data of the decedents was obtained from clinical record review. A DeCoDe panel evaluated the MITs-lite cases to determine the causal pathway to death, as undertaken for the CHAMPS study.

The Human Medical Human Research Ethics Committee of the University of Witwatersrand granted ethics consent for all studies and relevant hospital management approvals were obtained before study initiation. Written, informed consent was obtained from the mother/guardian.

### Study procedures

To prepare whole-genome sequencing libraries, stored isolates were sub-cultured, and genomic DNA extracted using standard methods. NexteraXT libraries were prepared (Illumina, San Diego, CA) and sequenced on the MiSeq platform generating 2 × 300 base paired-end (PE) reads (Illumina, San Diego, CA). Following sequencing, the quality of the raw reads was assessed using FASTQC v0.12.1 and processed using the Jekesa pipeline. Trim Galore removed adapters, ambiguous reads, and low-quality bases (Q >30 and length >50 bp). De novo assemblies were generated with SPAdes and Shovill, and QUAST 5 was used to evaluate their quality and to calculate assembly matrices. Sequences were uploaded onto the Pathogenwatch6 database for species confirmation and ConFindr was used to check for contamination. MLST and cgMLST predictions were performed using Klebsiella PasteurMLST and BIGSdb-Pasteur, respectively. Capsule K and O serotype predictions were made with Kaptive V3. Antimicrobial resistance genes identified with Kleborate compared to the curated version of the Comprehensive Antibiotic Resistance Database program including ResFinder 9.

Virulence genes were identified by Kleborate using the BLASTn for Key loci. Phylogenetic analysis was conducted using wgMLST and cgMLST. The sARGs were investigated through ResFinder. iTOL 12 was used to generate and visualize the phylogenetic tree.

### Outcomes

The primary objective of the study was to genetically characterize KPn isolates associated with invasive disease, including fatal cases, over six years in South African children, stratified by pHAI and presumed community-associated infections (pCAI). As an exploratory analysis, we analyzed temporal changes in Kpn isolates collected over six years, focusing primarily on isolates collected at the Chris Hani Baragwanath Academic Hospital (CHBAH), where most isolates were accrued.

### Statistical analysis

Multiple Kpn isolates collected from the same individual were sequenced only if collected more than 7 days apart, considering the high likelihood of the isolates being due to the same invasive disease episode. In cases where multiple isolates were sequenced from the same participant, only genetically distinct isolates were included. When KPn was cultured concurrently from different sites, sequencing was limited to a single isolate per participant, using a hierarchical approach of blood, CSF, and lung tissue. Early-onset invasive disease (EOD) was defined as culture-confirmed invasive KPn within 72 hours of birth, and late-onset disease (LOD) was defined as episodes diagnosed from 72 hours until ≤90 days of age. Further, pCAI were defined as invasive KPn detected on admission or within 72 hours of hospitalization or if the death occurred in the community [17]. Whereas, pHAI was defined as an invasive KPn detected more than 72 hours after admission to the hospital or if the DeCoDe panel attributed nosocomial infection to the causal pathway of the death. Multi-drug resistance (MDR) was defined as the presence of genetic markers in three or more antimicrobial classes.

Data was analyzed with Stata Version 11.0 (StataCorp, Texas, USA) and R Version 4.1.1 (Vienna, Austria). The findings from the four observational studies were aggregated in the main analysis. The distribution of the isolates stratified by each observational studies is shown in Supplementary Table 2. A multiple logistic regression model adjusting for possible covariates was used to compare the ST, K-loci, O antigen types, antimicrobial resistance markers (ARM), and virulence factors between pHAI and pCAI isolates. Percentages were reported alongside adjusted odds ratios (aOR) and 95% confidence intervals (CI). P-values of 0.05 were considered significant and no adjustment was undertaken for multiplicity in this hypothesis-generating study.

## RESULTS

Overall, 681 invasive disease Kpn isolates were identified during the four observational studies (435 identified during hospital surveillance and 246 from postmortem sampling); Table 1. Of those, 367 (53.9%) were sequenced. Reasons for sequencing not being undertaken included 84 isolates collected less than 7 days apart, 149 isolates not retrieved from the NHLS, and KPn not identified during sub-culture for 81 isolates. Thirteen additional isolates were excluded after sequencing as their genotypes were identical to an earlier isolate sequenced. Two decedents had a genomically identical KPn identified during antemortem and postmortem sampling, thus the postmortem isolates were excluded from further analysis. Lastly, 4.1% (15/367) of the isolates sequenced were identified as other Klebsiella species and excluded from further analysis. Two isolates from four different infants and three isolates from one infant, collected during hospital surveillance, were included in the final analysis as sequencing identified different genotypes.

**Table 1:** Study consort.

|  | **Total**<br>**number of**<br>**isolates** | **Observation Study 1:**<br>**Hospital surveillance**<br>**4 Mar 2019 - 27 Feb 2021** | **Observation Study 2:**<br>**Hospital surveillance**<br>**13 May 2022 - 31 Oct 2023** | **Observation Study 3:**<br>**Postmortem sampling - CHAMPS**<br>**17 Jan 17 - 19 Ap 2023** | **Observation Study 4:**<br>**Postmortem sampling - MITs lite**<br>**21 Feb 2018 - 15 Sept 2022** |
| --- | --- | --- | --- | --- | --- |
| ***K. Pneumoniae* cultures** | 681 | 217 | 218 | 136 <sup>#</sup> | 110 <sup>#</sup> |
| Collected <7 days apart | 84 | 44 | 40 | - | - |
| Case plate not retrieved from NHLS | 149 | 62 | 25 | 42 | 20 |
| *K. Pneumoniae* grown during sub-culture | 81 | 6 | 9 | 27 | 39 |
| **Cultures available for WGS** | 367 | 105 | 144 | 67 | 51 |
| Same genome as first * | 13 | 7 | 6 | - | - |
| Other Klebsiella species | 15 | 2 | 8 | 1 | 4 |
| Same genome was collected during hospital surveillance | 2 | - | - | 2 | 0 |
| **Cultures included in the final analysis** | 337 | 96 | 130 | 64 | 47 |

Abbreviations: CHAMPS, Child Health and Mortality Prevention Surveillance; MITs, Minimally invasive tissue sampling; NHLS, National Health Laboratory service.
\* Interval of isolates (days): 8, 10, 12, 16, 21, 22, 23, 24, 32, 39.
<sup>#</sup> *K. pneumoniae* isolates from decedents in which the organism was attributed to the causal pathway of death.

### Clinical and demographic characteristics

Of the 337 isolates included in the final analysis, 1.9 % (7/337) were from stillbirths, 4.2% (14/337) from neonates <72 hours of age, 22.8% (77/337) from neonates 3 to ≤7 days of age, 44.8% (151/337) from neonates 7 to ≤28 days of age, and 26.4% (89/337) from infants 28 to ≤90 days of age. Fifty-seven percent (191/337) of the isolates were from males, while 28.8% (97/337) were collected from cases born to women living with HIV (i.e. HIV-exposed; Supplementary Table 3). Overall, 70.0% (236/337), 11.6% (39/337), and 9.5% (32/337) of the sequenced isolates were cultured from blood, lung tissue, and CSF, respectively. Seventy-two percent (243/337) of the isolates were from infants born prematurely (<37 weeks gestation), while 82% (275/337) of the isolates were from pHAI episodes; Supplementary Table 3.

### Genomic diversity and phylogenetic analysis

There was a large diversity among KPn clones detected, with the majority (90.6%, 77/85) of clonotypes each contributing individually to less than 3% of the overall isolates; Figure 1a. In total, 48 K-loci and 7 O-antigens, with 85 combinations, were identified among the 70 ST genomes. The most prevalent clonotype identified was ST17 associated with K-locus (KL) 25 and the O5 (19.3%, 65/337) antigen, followed by ST39 with KL149 and O1ab (9.8%, 33/377). Phylogenetic analysis showed historical branching and descent from common ancestral strains (Figure 1b). There are three major clades in the tree, with the major clade comprising isolates associated with fatal outcomes, suggesting that these isolates might be genetically closer (i.e. have a common lineage) compared to those forming the other clusters. The three major clades were clustered based on the O antigen and K Locus with O5, O1ab, and O2afg clustering with KL25, KL149, and KL102 respectively. *K. pneumoniae* was identified in two sets of twins. The first twin dyad (66010-1 & 66101-2) had clades with different O antigens and K loci, albeit both being pHAI episodes. The second twin dyad (68936-1 and 68936-2) also had pHAI, with one twin forming part of the major clade, and the other subclade sharing the same O-antigen, but different K-loci, indicative of a different ancestorial history.

**Figure 1:**
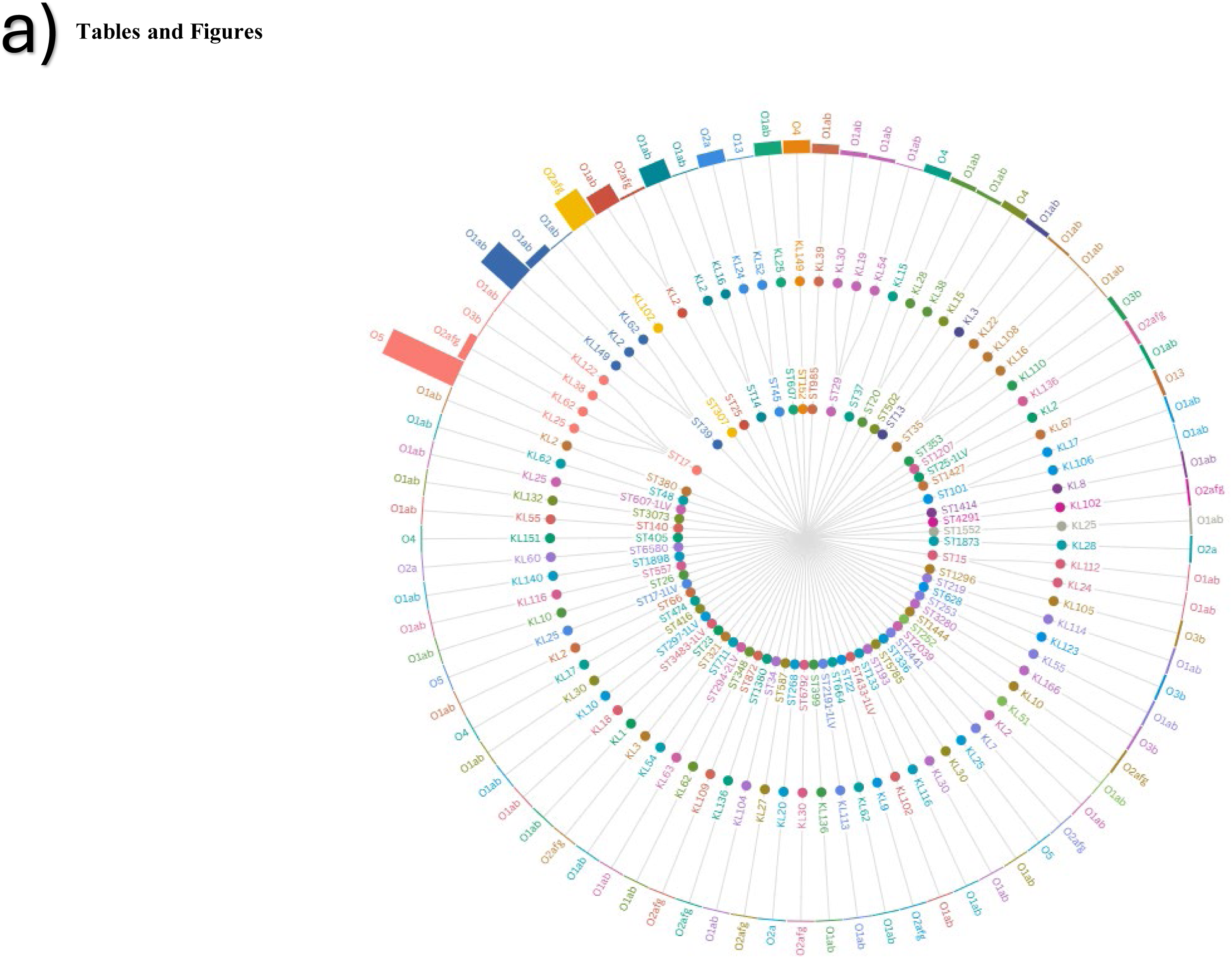
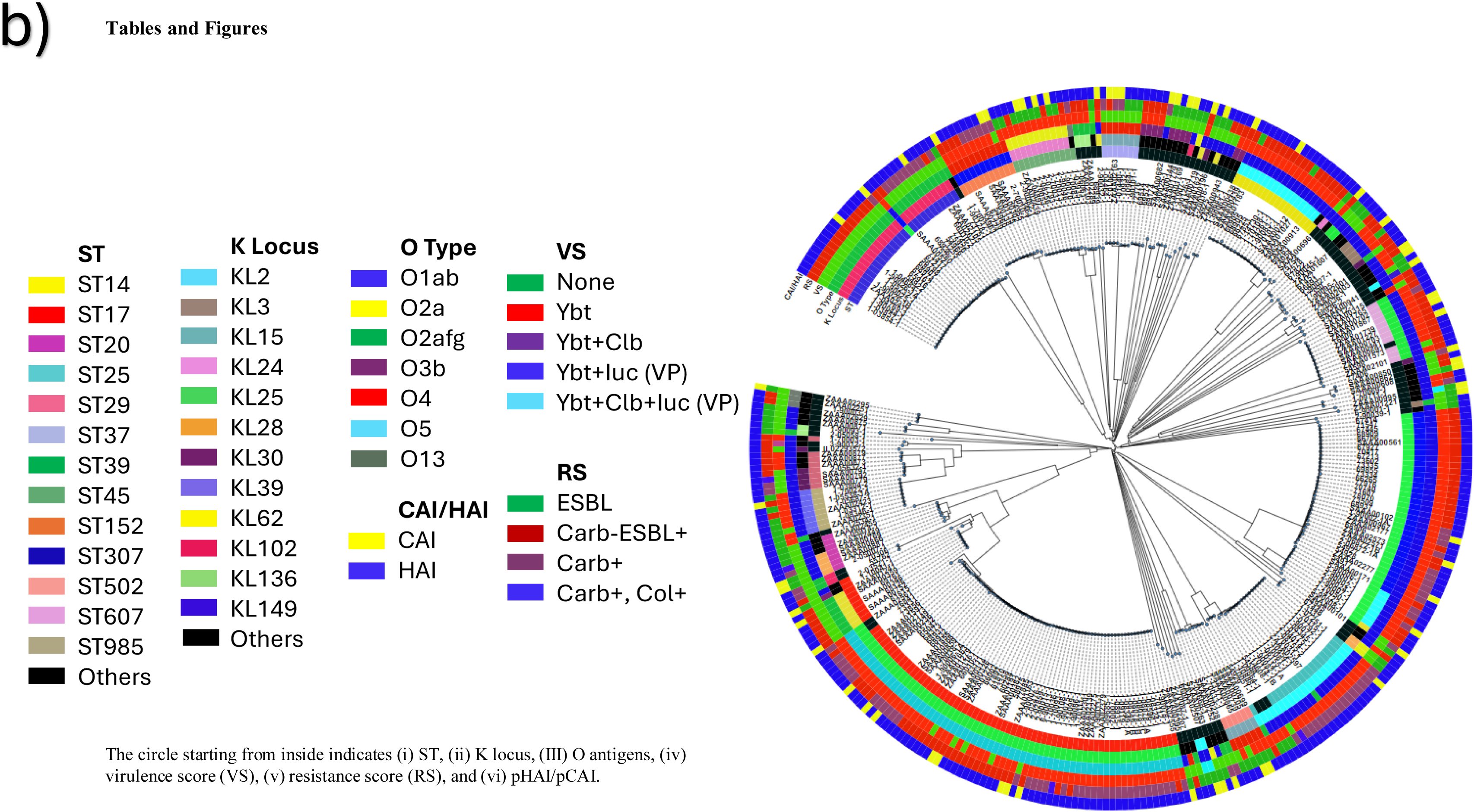
Hierarchical radical tree depicting the linkage between sequence types, K-loci, and O-antigens (a) and Phylogenetic tree of *K. pneumoniae* (b). Abbreviations: CAI, community-acquired; Clb, Colibactin; Carb, carbapenems; Col, colistin; ESBL, Extended Spectrum β-lactamase; HAI, hospital-associated; iuc, Aerobactin; RS, resistance score; ST, Sequence type; VS, virulence score; ybt, Yersiniabactin. HAI was defined as an invasive *K. pneumoniae* detected more than 72 hours after admission to the hospital or if the DeCoDe panel attributed nosocomial infection to the causal pathway of the death. CAI isolates were defined as invasive KPn detected on admission or within 72 hours of hospitalization or if the death occurred in the community. ST Others: ST8, ST13, ST15, ST16, ST18, ST20, ST23, ST37, ST48, ST73, ST111, ST140, ST252, ST253, ST297, ST299, ST327, ST348, ST416, ST530, ST551, ST610, ST789, ST985, ST0248, ST1401, ST1418, ST1427, ST1552, ST1728, ST1777, ST2534, ST2621, ST2938, ST2b07, ST3157, ST3280, ST4060, ST4845, ST4960, ST6078, ST7c08, ST92b1, and STf4da. K locus others: KL1, KL3, KL7, K12, KL13, KL14, KL16, KL20, KL24, KL39, KL48, KL49, KL60, KL63, KL64, KL67, KL106, KL111, KL112, KL113, KL140, KL143, KL158, and KL166 Figure one consists of two parts: (a) A hierarchical tree that shows the relationship between different Sequence types (ST), K-loci, and O-antigens in *Klebsiella pneumoniae*. (b). A phylogenetic tree representing the genetic relationship between the different *Klebsiella pneumoniae* strains identified in the study.

The distribution of the Kpn strains causing invasive disease collected from CHBAH varied across the six years (Figure 2). While ST17 isolates harbouring the KL25 capsule and O5 antigen were detected throughout the study period, ST39-KL149-O1ab (80.8%, 21/26) and ST307-KL102-O2afg (52.0%, 13/25) clonotypes were mainly detected between October 2019 and June 2020, respectively. The distribution of clonotypes, ST, K-loci, and O-antigens at the different collection sites are detailed in the Supplementary text and Supplementary Figure 1.

**Figure 2:**
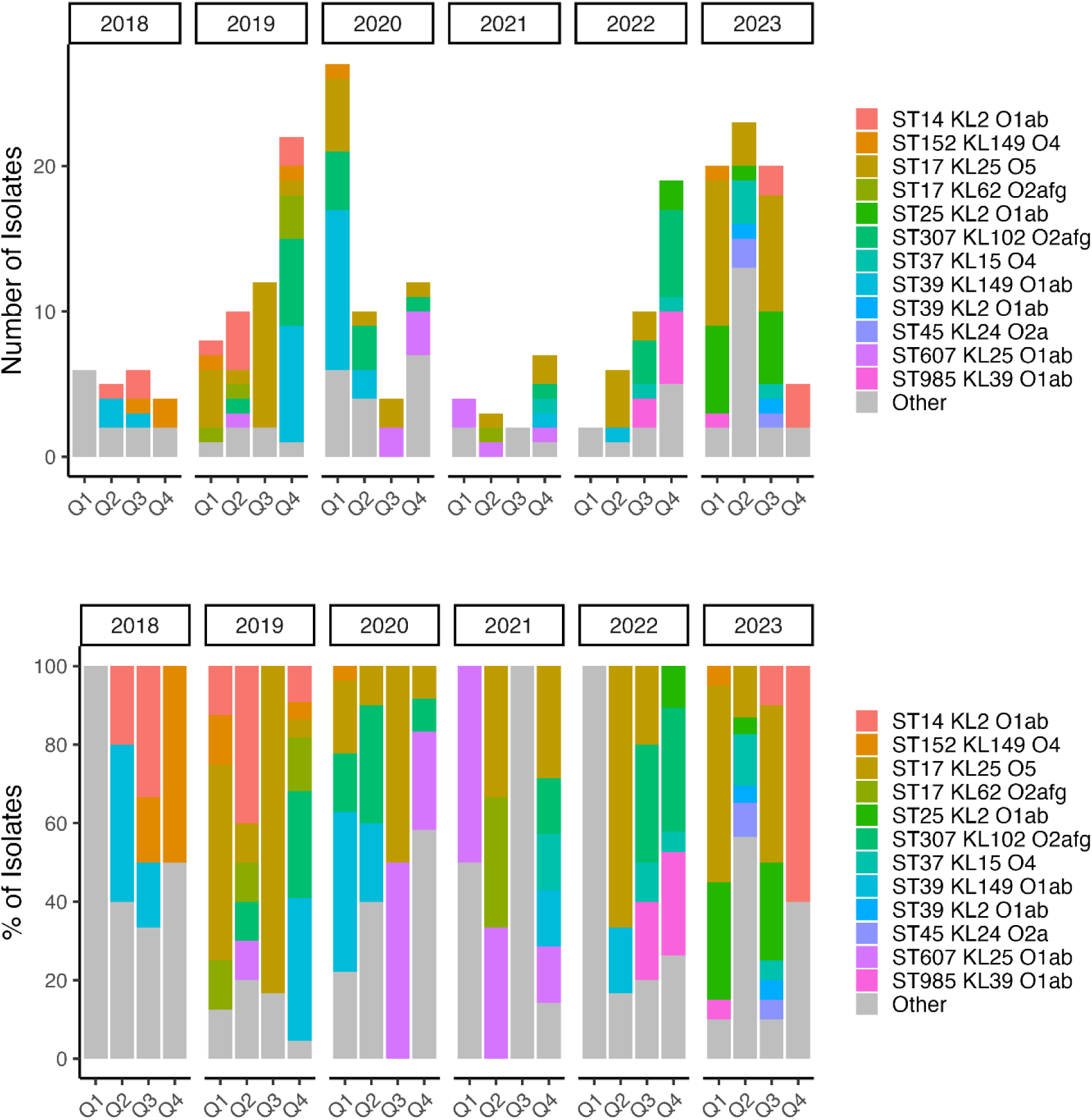
Changes in *K. Pneumoniae* genotypes causing invasive disease in South African infants*. \**Analysis limited to isolates collected at the Chris Hani Baragwanath hospital.* Graph showing the different *Klebsiella pneumoniae* strains that caused disease in young South African infants over six years.

### Sequence types (ST), Capsular polysaccharide (K-) Loci, and Lipopolysaccharide (O-) antigens

Overall, ST17 predominated (22.0%, 74/337), followed by ST39 (12.5%, 42/337; Table 2). There were no statistical differences in the prevalence of the STs between pHAI and pCAI isolates; however, there was a greater diversity of STs in pHAI (n=46) than in pCAI (n=21) isolates; Supplementary Figure 2a. Overall, KL25 predominated (24.0%, 81/337), followed by KL2 (14.5%; 49/337), KL149 (13.4%, 45/337), and KL102 (9.5%, 32/337; Table 2). The dominant O-antigen detected was O1ab (48.1%, 162/337) followed by O5 (19.9%, 67/337; Table 2). The prevalence of the K-loci and O-antigens were similar between pCAI and pHAI isolates, except for a lower prevalence of KL62 (2.2%, 6/275 vs. 8.1%, 5/62; p=0.034) and O3b in pHAI isolates (2.2%, 6/275 vs. 8.1%, 5/62; p=0.027), respectively; Supplementary Figure 2b-c.

**Table 2:**
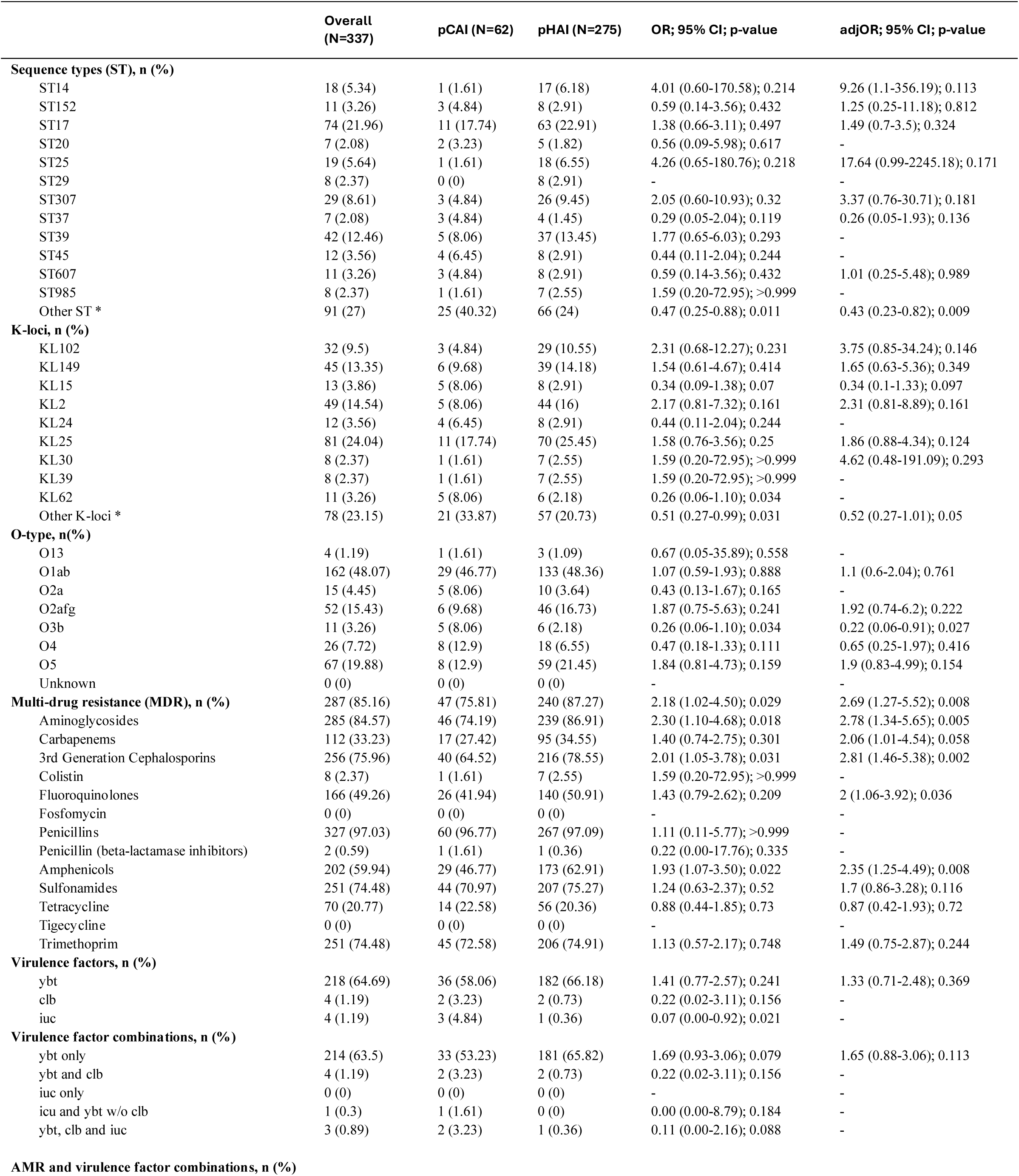
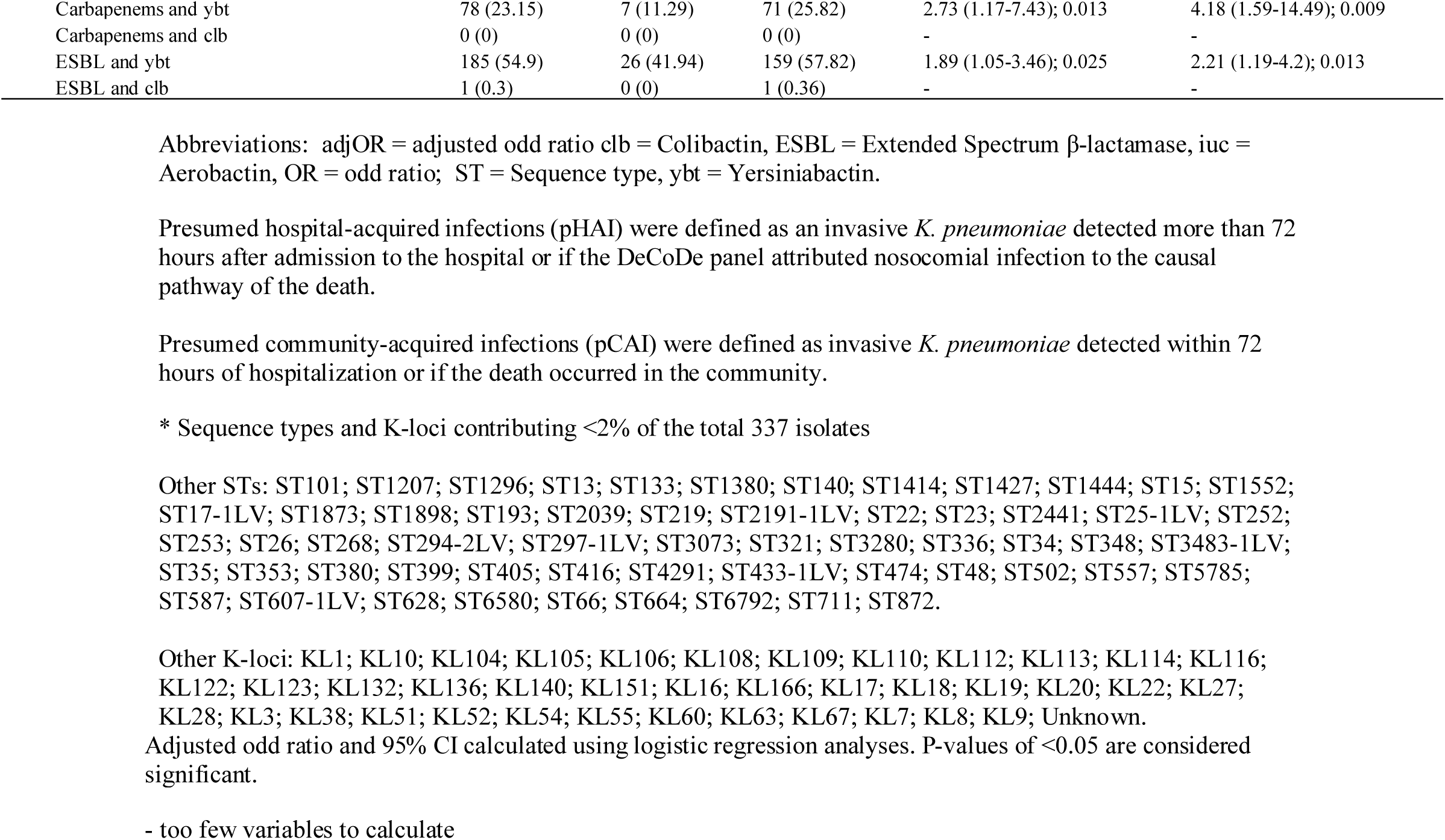
Genomic characterization of *K. pneumoniae* causing invasive disease and mortality in South African infants 0-90 days of age.

The prevalence of ST, K-loci, and O-antigens stratified by collection site is detailed in Supplementary Table 4.

### Antimicrobial resistance

Overall, 85.2% (287/337) of the KPn isolates harboured MDR genes, including 84.6% (285/337) harbouring genes encoding resistance to aminoglycosides, 76.0% (256/337) to 3^rd^ generation cephalosporins, 33.2% (112/337) to carbapenems, and 2.4% (8/337) to colistin (Table 2). There was a 2.69 higher odds of MDR genes in pHAI (87.3%, 240/275) compared with pCAI (75.8%, 47/62; p=0.008) isolates, including a higher prevalence of genes conferring resistance to aminoglycosides (86.9%, 239/275 vs. 74.1%, 46/62; p=0.005) and 3^rd^ generation cephalosporins (78.6%, 216/275 vs. 64.5%, 40/62; p=0.002; Table 2).

Overall, the TEM-1D (bla_TEM-1D_) and CTX-M-15 (bla_CTX-M-15_) β-lactamase genes were detected in 61.4% (207/337) and 75.1% (253/337) of the isolates, respectively whilst the oxacillinase-type β-lactamase-48 (bla_OXA-181_) gene was detected in 26.4% (89/337) of the isolates. The New Delhi metallo β-lactamase-5 (bla_NDM-5_) and New Delhi metallo β-lactamase-1 (bla_NDM-1_) genes were detected in 2.1% (7/337) and 0.3% (1/337) of the isolates, respectively (Supplementary Table 5). The prevalence of the antimicrobial resistance genes was similar between pHAI and pCAI isolates except for a higher prevalence of bla_TEM-1D_ (64.4%, 177/275 vs. 48.4%, 30/62; p=0.021) and bla_CTX-M-15_ (77.8%, 214/275 vs. 62.9%, 39/62; p=0.022) in pHAI compared with pCAI, respectively.

Among the strains with more than 6 isolates, ST152, ST25, ST307, and ST37 isolates were associated with the highest (median: 6 classes each) and ST20 (median: 1 class) the lowest number of antimicrobial resistant (AMR) classes; Supplementary Figure 3a. The resistance scores, which identify clonotypes warranting escalation of antimicrobial therapy [18], varied by ST; Figure 3a. The prevalence of genes encoding resistance to Extended-Spectrum Beta-Lactamases (ESBL) production was highest in ST607 (90.9%, 10/11) and lowest in ST20 (0%, 0/7) and ST502 (0%, 0/6) isolates. The prevalence of genes encoding resistance to carbapenem was highest in ST1444 (100%, 2/2) and ST25 (94.7%, 18/19) isolates - including a single ST25 isolate that also harboured genes encoding for colistin resistance, and lowest in ST14 (0%, 0/18) and ST985 (0%, 0/8) isolates.

**Figure 3:**
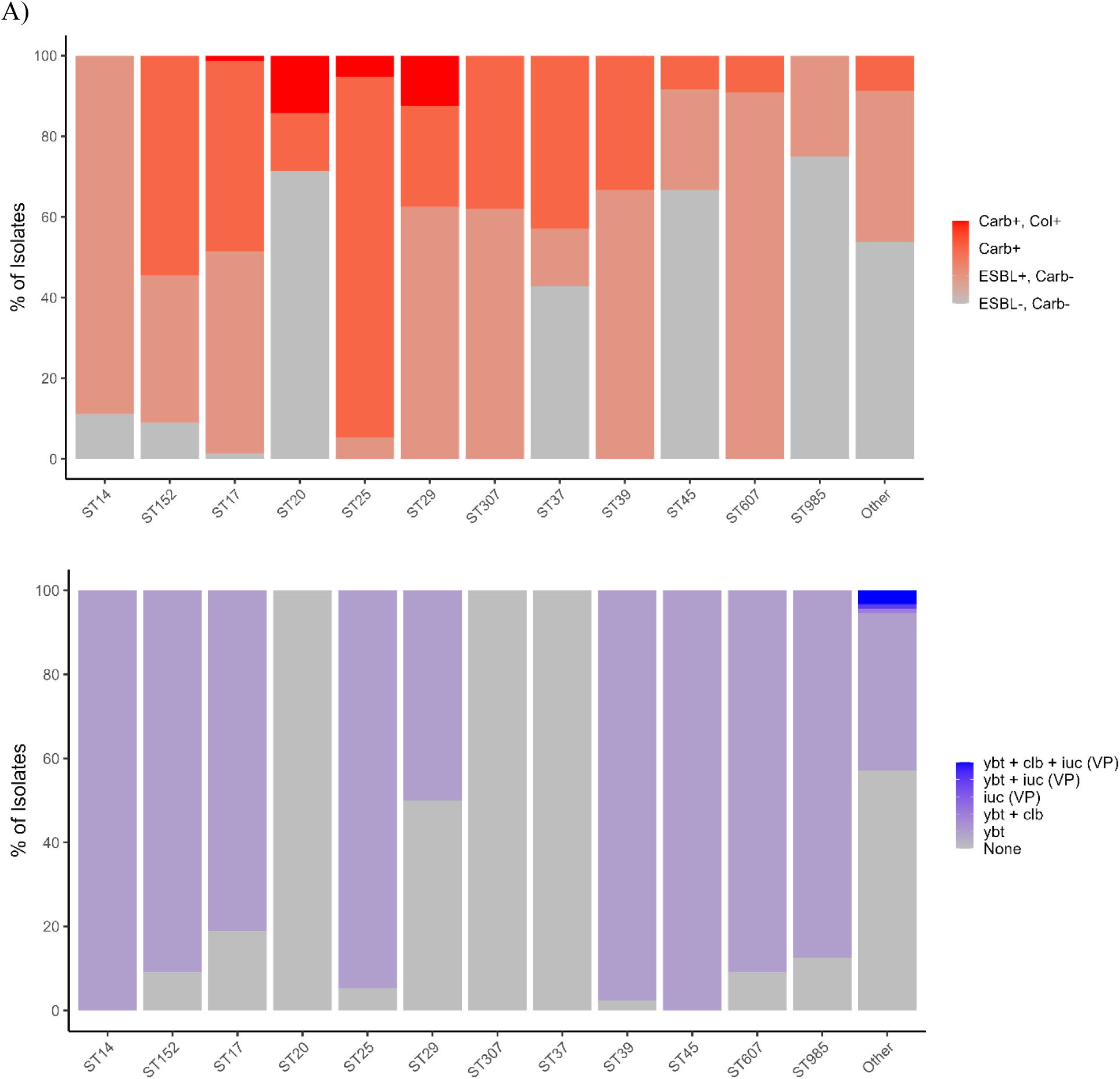
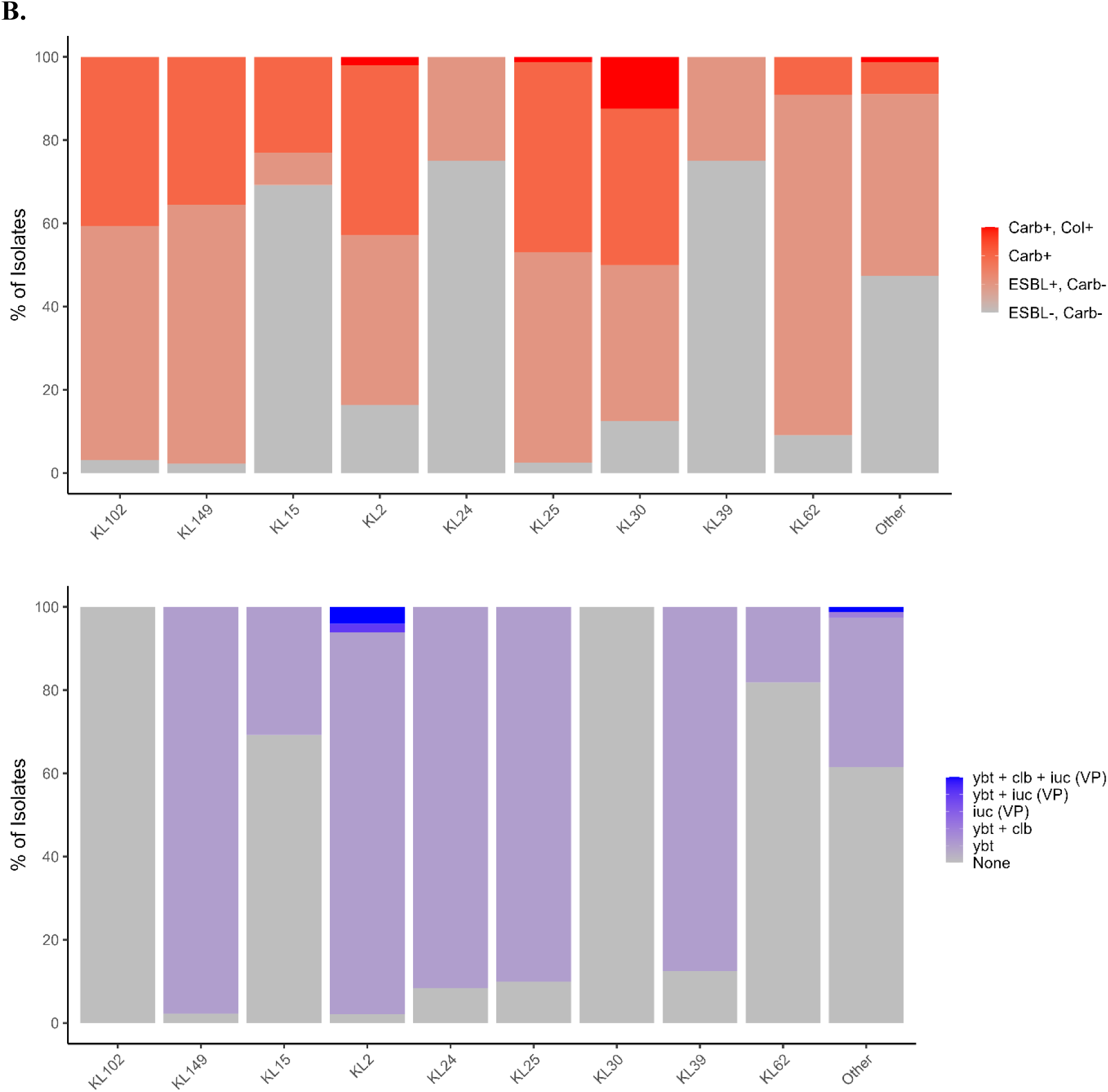
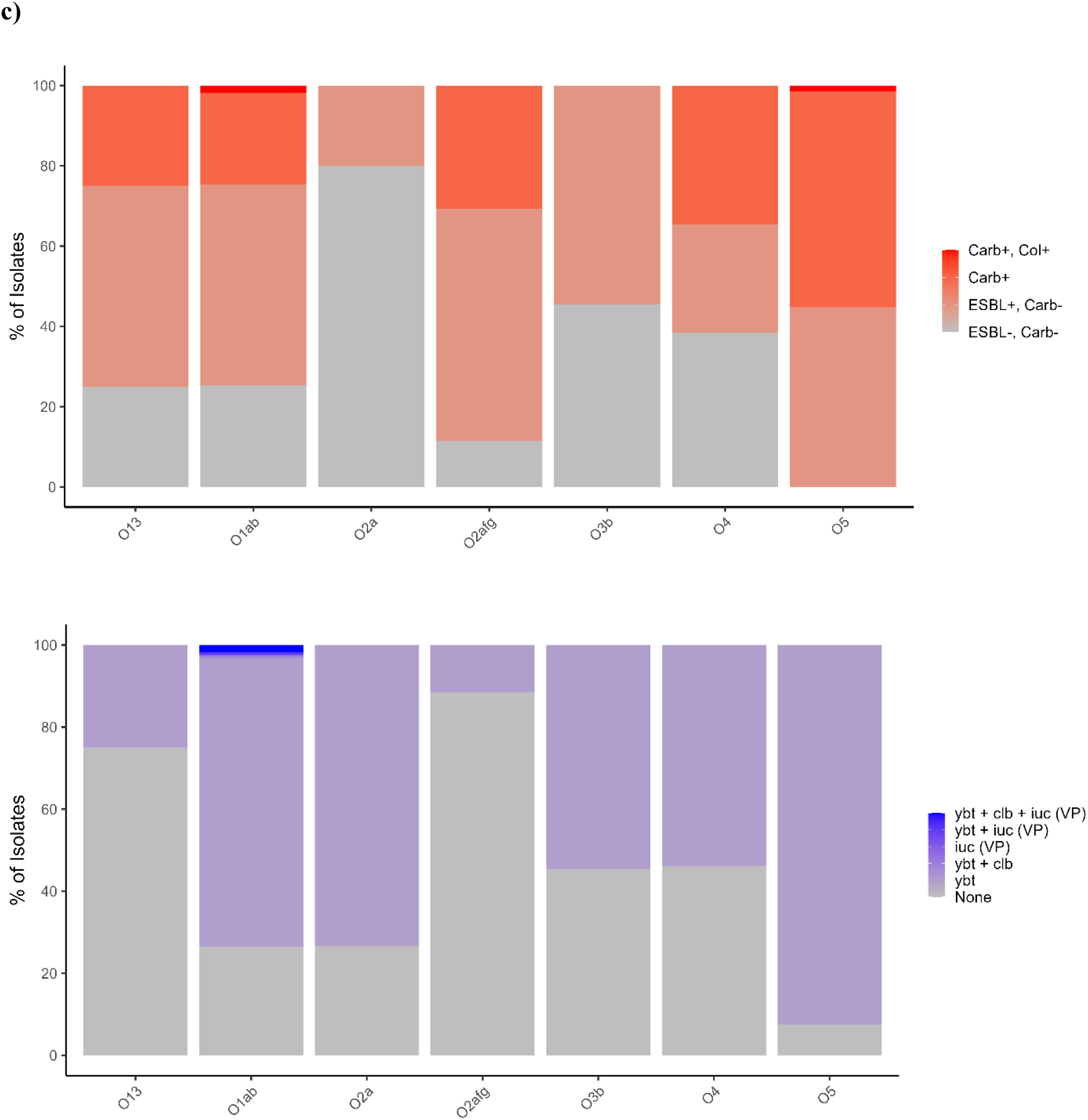
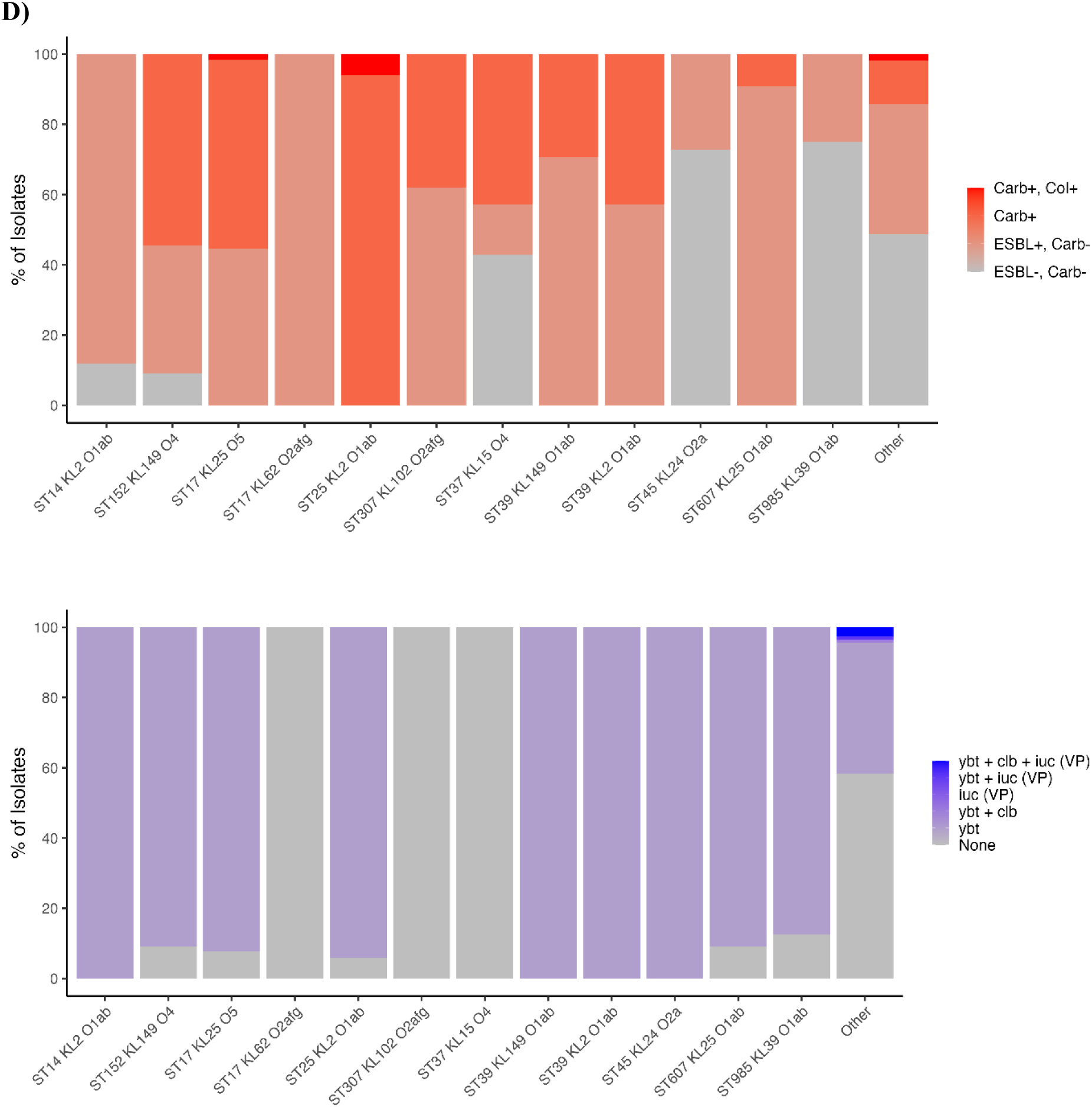
Distribution of resistance and virulence scores, stratified by common ST (a), K-loci (b), and O- antigens (c) and clonotypes (d). Abbreviations: Clb, Colibactin; Carb, carbapenems; Col, colistin; ESBL, Extended Spectrum β-lactamase; iuc, Aerobactin; ST, Sequence type; ybt, Yersiniabactin. Graphs showing the distribution of resistance (in red) and virulence scores (in purple) of *Klebsiella pneumoniae* strains detected in the study across different categories. Panel (a) displays the distribution stratified by the common Sequence types (ST). Panel (b) shows the distribution based on the K-loci, while panel (c) represents the distribution according to the O-antigens. Lastly, panel (d) presents the distribution based on clonal types.

The KL102 and KL30 isolates were associated with the highest number of AMR classes (median: 6 classes each), whereas KL62, KL39, KL15, and KL149 encoded resistance to fewer classes (median: 3 classes each); Supplementary Figure 3b. The prevalence of genes encoding for ESBL production was highest in KL62 (81.8%, 9/11) and lowest in KL15 (7.7%, 1/13); Figure 3b. The prevalence of genes encoding resistance to carbapenem varied by K-loci, with the highest prevalence detected in KL25 (51.8%, 42/81). Two KL25 (2.5%; 2/81), two KL2 (4.1%;2/49), and a single KL30 (12.5%; 1/8) isolate, respectively harboured genes encoding resistance to both carbapenem and colistin; Figure 3b.

The O2afg, O4, and O5 isolates were associated with the highest number of AMR classes (Median: 6 classes each), whereas O1ab encoded resistance to fewer classes (median: 4 classes); Supplementary Figure 3c. The prevalence of genes encoding for ESBL production was highest in O2afg (57.7%, 30/52) and lowest in O2a (15%, 3/15) isolates; Figure 3c. The prevalence of genes encoding carbapenem resistance varied by O antigen (0-61.2%) with the highest prevalence detected in O5 (61.2%, 41/67).

When stratifying by the most identified clonotypes, all ST25-KL2-O1ab isolates (n=17) harboured genes associated with carbapenem resistance, including a single isolate that also harboured genes for colistin resistance; Figure 3d. All ST307-KL102-O2afg (62.1%, 18/29 and 37.9%, 11/29), ST39-KL149-O1ab (70.6%, 24/34 and 29.4%, 10/34), ST607-KL25-O1ab (90.9%, 10/11 and 9.1%, 9/11), and ST17-KL25-O5 (44.6%, 29/65 and 55.4%, 36/65) genotypes harboured genes for ESBL production or carbapenems resistance, respectively, including a single ST17-KL25-O5 isolate that harboured genes encoding for both carbapenems and colistin resistance. Lastly, most of the ST152-KL149-O4 genotypes harboured genes encoding for ESBL production (54.5%, 6/11) or carbapenems (36.4%, 4/11) resistance, while no resistance genes were detected in a single ST152-KL149-O4 isolate.

### Virulence factors

Yersiniabactin (ybt), associated with hypervirulence, was the dominant virulence factor detected in 64.7% (218/337) of the isolates, while genes encoding for colibactin (clb, 4/337) and aerobactin (iuc, 4/337) virulence were detected in <2% of isolates; Table 2. Twenty-three percent (78/337) of the isolates harboured genes encoding for ybt virulence and carbapenems resistance. There was a 4.18 higher odds of harbouring genes encoding for both ybt virulence and carbapenems resistance in pHAI (25.8%, 71/275) compared with pCAI (11.3%, 7/62; p=0.009) isolates. (Table 2). Fifty-five percent (185/337) of isolates harboured genes encoding for both ybt and ESBL production, with a higher prevalence in pHAI (57.8%, 159/275) compared with pCAI (41.9%, 26/62; aOR 2.21; 95% CI:1.19-4.2 p=0.013) isolates. The distribution of virulence factors stratified by clonotypes, ST, K-loci, and O antigens is illustrated in Figures 3a-d

## DISCUSSION

We documented a wide diversity of MDR KPn strains associated with invasive disease in South African infants up to 90 days of age between 2018 and 2023. We also identified several high-risk clonotypes, including ST25- KL2-O1ab, with all isolates harbouring genes associated with carbapenem resistance. Furthermore, the ST307- KL102-02afg, ST39-KL149-01ab, ST607-KL25-O1ab, ST17-KL25-O5, and ST152-KL149-O4 clonotypes were associated with a high prevalence of genes encoding for ESBL production or carbapenem resistance. Most ST25, ST607, ST307, ST39, ST152, and ST17 clones also harboured genes for Yersiniabactin (81-100%) virulence. While the ST25, ST17, ST307, and ST39 clones have been reported elsewhere, including South Africa [19–21], the ST607 clones are rare and have only been reported in a limited number of invasive isolates from China and France [22].

Overall, most of the KPn isolates sequenced harbored MDR genes, spanning resistance to first, second, and third-line therapeutic options. This aligns with global reports of increased acquisition of antibiotic resistance and the broader antimicrobial resistance landscape in South Africa and poses significant challenges for managing KPn infections in our setting [23, 24]. The emergence of MDR and highly virulent strains harbouring genes for ybt virulence along with ESBL production or carbapenems resistance is particularly worrying in terms of the clinical ramifications of increased severity of infections, limited treatment options, increased case fatality risk, and potential for rapid dissemination within healthcare facilities [25, 26]. In South Africa, the standard treatment for young infants suspected of having community-acquired sepsis includes ampicillin and gentamicin treatment while pHAI is treated with third-generation cephalosporins or carbapenems. The AMR profile of the sequenced Kpn strains in this study indicates that these treatment options could largely be ineffective, underscoring the critical need for vaccines or alternative, more effective drug combinations.

Although the prevalence of ST, K-loci, and O-antigens was similar between pHAI and pCAI isolates, pHAI isolates had a 2.69 higher odd of harbouring MDR genes and a 4.18 higher odd of harbouring genes encoding for both ybt virulence and carbapenems resistance compared with pCAI isolates. The heightened prevalence of MDR among isolates acquired within healthcare settings emphasizes the significant role that healthcare facilities play in the spread of antimicrobial resistance. It is, however, noteworthy that seventy-six percent of the pCAI isolates sequenced also harboured MDR genes which may be attributed to high exposure to antibiotics in our setting or dissemination of pHAI strains in the community.

Studies are underway on developing a vaccine to protect against the most common Kpn strains; however, the geographic variability in strain distribution poses a significant challenge to vaccine development [27]. In our setting, a vaccine achieving over 80% coverage would need to target eleven capsular serotypes to protect against invasive disease. The dominant capsular serotypes identified in our setting included KL25, KL149, KL2, KL102, and KL15, which largely align with global distributions, except for KL149, reported only in a few studies from Africa [28–30] and Poland [31]. The major K-loci detected in our setting and globally are important targets for consideration for multivalent K-antigen vaccine development. Nevertheless, variations in KPn lineages observed in this study and over 20 years in Malawi [32] underscore the need for ongoing surveillance if a multi-valent K-antigen based vaccine is developed, considering the fluctuation of dominant lineages over time and the potential of serotype replacement disease by non-vaccine serotypes. A vaccine targeting the lipopolysaccharide serotypes, which are fewer in number and remained more stable throughout the study period, may be less prone to replacement disease by non-vaccine serotypes, with the inclusion of three O-antigens (O1ab, O2afg, and O5) covering over 80% of strains in our study.

Limited studies have characterized colonizing KPN isolates in Africa, including South Africa, primarily due to challenges in surveillance and diagnostic infrastructure. Nevertheless, a previous study by our group examining the genomic relatedness between a limited number of colonizing and invasive Kpn strains in South African infants indicated that there might be some genomic differences between these groups [33]. The observations included that invasive compared with colonizing strains had a narrower range of lineages, higher prevalence of multidrug resistance (MDR) genes, and higher prevalence of yersiniabactin; however, further research is needed to fully elucidate the relationship between colonizing and invasive strains, particularly in African settings.

Strengths of this study included that isolates of KPn were identified through both observational surveillance and post-mortem tissue sampling and were not biased towards selecting isolates associated with MDR. Moreover, isolates were collected from young infants (<90 days of age) with most other studies focusing on older children and adults. Lastly, a large number of isolates were sequenced with appropriate associated clinical and demographic data. Limitations of this study included that most of the KPn strains sequenced were identified at a single site (CHBAH), limiting the generalizability of the data to a national level. Further, there is some uncertainty in the definitions used to classify infections, as pCAI and pHAI which may not conclusively indicate the source of infection. The sample size for pCAI isolates was small, thus results should be interpreted with caution. Also, phenotypic susceptibility results were unavailable and could not be cross-checked against the genotypic ARM data. This study was hypothesis generating and not powered to investigate individual differences in ST, K-loci, and O-antigen, and we thus did not adjust for multiplicity. Lastly, antemortem sampling alone, particularly in settings with high case fatality rates, has low sensitivity compared to combined ante-mortem and post-mortem sampling, where multiple sites are sampled and not restricted by low blood volume. Consequently, data from the four observational studies were aggregated for the main analysis; however, this may have introduced some bias weighted towards more virulent Kpn strains or those not covered by standard empirical antimicrobial regimens and findings should be interpreted with caution.In conclusion, although there was a wide diversity of strains associated with Kpn invasive disease in South African children, over 80% of the isolates were attributed to eleven capsular loci or three lipopolysaccharide (O-antigen) serotypes. The findings from this study could be useful in selecting KPn antigen targets for potential vaccine candidates. Moreover, the changing epidemiology of MDR KPn causing invasive disease in South African infants emphasizes the need for continuous genomic characterization in LMIC regions to track changes in virulence and antibiotic susceptibility profiles.

## Data Availability

The raw PE reads will be available through GenBank upon acceptance of publication. Associated metadata will be made available to researchers who provide a methodologically sound proposal. Data will only be made available if approval is granted from the Human Research Ethics Committee, University of Witwatersrand, Johannesburg, South Africa. Further, all requesters will need to sign a data transfer agreement.

## Authors contributions

CPO and SAM conceived and designed the study, and secured funding. SJ, CB, GK, VLB, SM, KS, ID, FLN, SCV, JW, RS, AMVN, NN, YR, MS, ZD contributed material. Sequencing data was generated by LVDM, NJD, LA, and UR under the supervision of RGKD, RS, and CPO. AI conducted the statistical analysis, while SK performed the bioinformatics analysis. CPO drafted the manuscript and all authors contributed to subsequent drafts, read, and approved the final version of the manuscript. CPO, AI, and SK verified the underlying data of this study. All authors had access to the data used in this study and accepted responsibility for the submission.

## Data availability

The raw PE reads will be available through GenBank upon acceptance of publication.

## Acknowledgments

We thank the infants who participated in the study and their parents. The views expressed in the publication are those of the author(s) and not necessarily those of the University of Witwatersrand, the National Research Foundation, or the Bill and Melinda Gates Foundation.

