## Supplementary for "Genomic characterization of *Klebsiella pneumoniae* causing invasive disease in South African infants: observational studies between 2018 to 2023"

#### SUPPLEMENTARY TEXT

##### ***Study population***

*K. pneumoniae* (KPn) isolates were collected from a sero-epidemiological study that aimed to identify serological markers associated with risk reduction of invasive Group B *streptococcus* (GBS) disease in infants less than or equal to 90 days of age [1]. Briefly, the study was undertaken from March 4th, 2019 to February 27th, 2021, and included a longitudinal, prospective cohort of 17 752 mother-newborn dyads who were enrolled at the Chris Hani Baragwanath Academic Hospital (CHBAH) and the Rahima Moosa Mother and Child Hospital (RMMCH) in the city of Johannesburg, South Africa. Women were enrolled either antenatally, during the early stages of labour, or immediately postpartum, and the infants born to the women in the cohort were followed up to 90 days of age for all-cause hospitalization and death. In addition to the infants enrolled as part of the cohort, additional infants who were not part of the cohort but identified through daily laboratory-based surveillance at the National Health Laboratory Service (NHLS) as cases of invasive *K. pneumoniae* disease, where *K. pneumoniae* was isolated from a normally sterile site (blood, CSF, joint fluid, pleural fluid) were also enrolled at the two facilities where the cohort cases had been enrolled and at four other hospitals in South Africa. The additional hospitals where enrolment of invasive *K. pneumoniae* cases was undertaken, following identification thereof through laboratory-based surveillance were at the Charlotte Maxeke Johannesburg Academic Hospital (CMJAH) in the City of Johannesburg, the Tshwane Academic Laboratory Network (TALM), Prince Mshiyeni Memorial Hospital (PMMH) in eThekwini and Mowbray Maternity Hospital (MMH) Cape Town. Surveillance for invasive *K. pneumoniae* isolates and enrolment of cases continued across all six sites until February 27th, 2021.

We also analyzed *K. pneumoniae* isolates collected from infants up to 90 days of age who were enrolled in a second hospital surveillance study that aimed to strengthen the evidence base of immunological correlates of protection against invasive GBS disease. Briefly, a cohort of 21,202 mother-infant dyads were enrolled between May 22nd 2022 to December 31st, 2023 at the CHBAH or RMMCH and infants were followed up to 90 days of life for all-cause hospitalization and death (cohort cases). Infants who were not part of the cohort (non-cohort cases), but were identified through daily laboratory-based surveillance of the same six sites as the first hospital surveillance study.

##### ***Temporal variation of sequence types (ST) over the study period***

The distribution of the ST causing invasive disease at CHBAH varied across the six years; Supplementary Figure 1. While ST17 was detected throughout the study period, ST39 and ST307 were mainly detected between October 2019 and May 2020 (75%, 21/28, and 52%, 13/28, respectively). Furthermore, all the ST25 (n=25) isolates were detected only between October 2022 and September 2023. The distribution of ST observed at the other collection sites is shown in Supplementary Figure 1.

***Temporal variation of k-loci over the study period***

The distribution of K- loci at CHBAH varied over the study period, with 69.7% (23/33) of the KL149 isolates identified between October 2019 and June 2020, 67.5% (25/37) of KL2 isolates detected between October 2022 and December 2023, and 50% (6/12) of KL15 detected between January and September 2023; Supplementary Figure 1. While KL25 isolates were not detected in 2018, they were detected throughout the study period from January 2019. Lastly, 50% (14/28) of the KL102 isolates were detected between October 2019 and June 2020 (53.6%, 15/28), while 32.1% (9/28) were detected between July and December 2022. The distribution of K-loci observed at the other collection sites is shown in Supplementary Figure 1.

***Temporal variation of O-antigen over the study period***

Although O1ab isolates were detected throughout the study period at CHBAH, a higher number of isolates were detected between January and December 2020 (26.1%, 30/115) and between January and December 2023 (25.2%, 29/115); Supplementary Figure 1. Moreover, 45.5% (25/55) of the O5 isolates were detected between January 2019 and December 2020 and 38.1% (21/55) between January and September 2023. While the O2afg and O4 isolates were detected throughout the study period, 75% (3/4) of the O3b isolates were detected between April and June 2023. Moreover, all of the O3b (n=3) isolates were detected between January and December 2022, while 57.1% (4/7) of the O2a isolates were detected between April and December 2023. The distribution of O antigens observed at the other collection sites is shown in Supplementary Figure 1.

### SUPPLEMENTARY TABLES

**Supplementary Table 1: Comparison of study procedures undertaken in the CHAMPS and Mits-lite studies.**

| Category | | CHAMPS | MITs Lite |
|---|---|---|---|
| Eligibility | | | |
| | Stillbirths | √ | |
| | Neonates and Paediatrics | √ | √ (only DOA's from 2021) |
| | CHAMPS catchment | √ | |
| | Greater Soweto | | √ |
| | Verbal autopsy | √ | √ (only from 2021) |
| Specimen collection | | | |
| | Blood culture | √ | √ |
| | CSF culture | √ | √ |
| | Lung culture | √ | √ |
| | HIV PCR | √ | √ |
| | NP swab | √ | √ |
| | Rectal swab | √ | √ |
| | Malaria testing | √ | |
| | Brain sample (occipital and transnasal) | √ | √ (only from 2021) |
| | Bilateral lungs | √ | √ |
| | Liver | √ | √ (only from 2021) |
| | Placenta for neonates and stillbirths | √ | |
| Testing | | | |
| | Histopathology | √ | √ |
| | Central pathology | √ | |
| | TAC | √ (done on TAC array) | √ (done on the open array) |
| | Lungs | √ | √ |
| | CSF | √ | Only done on dead on arrival from 2021 |
| | Rectal | √ | |
| | Covid PCR | √ | √ |

Abbreviations: CSF – cerebral spinal fluid; CHAMPS – Child Health and Mortality Prevention Surveillance; HIV – Human Immunodeficiency Virus; MITs – minimally invasive tissue sampling; NP – nasopharyngeal.

**Supplementary Table 2: Genomic characterization of *K. pneumoniae* causing invasive disease and mortality in South African infants 0-90 days of age, by observational study**

| | Observation Study 1: Hospital surveillance 4 Mar 2019 - 27 Feb 2021 | Observation Study 2: Hospital surveillance 30 April 2022 – 13 Nov 2023 | Observation Study 3: Postmortem sampling - CHAMPS 17 Jan 2017 - 19 Ap 2023 | Observation Study 4: Postmortem sampling - MITs lite 21 Feb 2018 - 15 Sept 2022 |
|---|---|---|---|---|
| **Sequence types, n(%)** | | | | |
| ST101 | 0/24 (0) | 0/74 (0) | 0/96 (0) | 1/5 (20) |
| ST1207 | 1/24 (4.17) | 1/74 (1.35) | 0/96 (0) | 0/5 (0) |
| ST1296 | 0/24 (0) | 2/74 (2.7) | 0/96 (0) | 0/5 (0) |
| ST13 | 0/24 (0) | 1/74 (1.35) | 3/96 (3.12) | 0/5 (0) |
| ST133 | 0/24 (0) | 1/74 (1.35) | 0/96 (0) | 0/5 (0) |
| ST1380 | 0/24 (0) | 1/74 (1.35) | 0/96 (0) | 0/5 (0) |
| ST140 | 0/24 (0) | 0/74 (0) | 0/96 (0) | 0/5 (0) |
| ST1414 | 0/24 (0) | 0/74 (0) | 1/96 (1.04) | 0/5 (0) |
| ST1427 | 1/24 (4.17) | 1/74 (1.35) | 0/96 (0) | 0/5 (0) |
| ST1444 | 0/24 (0) | 0/74 (0) | 0/96 (0) | 0/5 (0) |
| ST15 | 0/24 (0) | 0/74 (0) | 1/96 (1.04) | 0/5 (0) |
| ST1552 | 0/24 (0) | 0/74 (0) | 2/96 (2.08) | 0/5 (0) |
| ST17-1LV | 1/24 (4.17) | 0/74 (0) | 0/96 (0) | 0/5 (0) |
| ST1873 | 0/24 (0) | 0/74 (0) | 2/96 (2.08) | 0/5 (0) |
| ST1898 | 0/24 (0) | 0/74 (0) | 0/96 (0) | 0/5 (0) |
| ST193 | 0/24 (0) | 0/74 (0) | 1/96 (1.04) | 0/5 (0) |
| ST2039 | 0/24 (0) | 0/74 (0) | 1/96 (1.04) | 0/5 (0) |
| ST219 | 0/24 (0) | 0/74 (0) | 0/96 (0) | 0/5 (0) |
| ST2191-1LV | 0/24 (0) | 1/74 (1.35) | 0/96 (0) | 0/5 (0) |
| ST22 | 0/24 (0) | 0/74 (0) | 0/96 (0) | 0/5 (0) |
| ST23 | 0/24 (0) | 0/74 (0) | 0/96 (0) | 0/5 (0) |
| ST2441 | 0/24 (0) | 0/74 (0) | 1/96 (1.04) | 0/5 (0) |
| ST25-1LV | 0/24 (0) | 1/74 (1.35) | 0/96 (0) | 0/5 (0) |
| ST252 | 0/24 (0) | 0/74 (0) | 1/96 (1.04) | 0/5 (0) |
| ST253 | 0/24 (0) | 0/74 (0) | 0/96 (0) | 0/5 (0) |
| ST26 | 1/24 (4.17) | 0/74 (0) | 0/96 (0) | 0/5 (0) |
| ST268 | 0/24 (0) | 0/74 (0) | 0/96 (0) | 0/5 (0) |
| ST294-2LV | 0/24 (0) | 0/74 (0) | 0/96 (0) | 0/5 (0) |
| ST297-1LV | 0/24 (0) | 0/74 (0) | 0/96 (0) | 0/5 (0) |
| ST3073 | 0/24 (0) | 0/74 (0) | 0/96 (0) | 0/5 (0) |
| ST321 | 0/24 (0) | 0/74 (0) | 0/96 (0) | 0/5 (0) |
| ST3280 | 0/24 (0) | 0/74 (0) | 0/96 (0) | 0/5 (0) |
| ST336 | 0/24 (0) | 0/74 (0) | 1/96 (1.04) | 0/5 (0) |
| ST34 | 0/24 (0) | 1/74 (1.35) | 0/96 (0) | 0/5 (0) |
| ST348 | 0/24 (0) | 1/74 (1.35) | 0/96 (0) | 0/5 (0) |
| ST3483-1LV | 1/24 (4.17) | 0/74 (0) | 0/96 (0) | 0/5 (0) |
| ST35 | 0/24 (0) | 1/74 (1.35) | 1/96 (1.04) | 0/5 (0) |
| ST353 | 0/24 (0) | 0/74 (0) | 4/96 (4.17) | 0/5 (0) |
| ST380 | 0/24 (0) | 0/74 (0) | 0/96 (0) | 1/5 (20) |
| ST399 | 0/24 (0) | 0/74 (0) | 0/96 (0) | 0/5 (0) |
| ST405 | 0/24 (0) | 0/74 (0) | 0/96 (0) | 0/5 (0) |
| ST416 | 0/24 (0) | 0/74 (0) | 0/96 (0) | 0/5 (0) |
| ST4291 | 0/24 (0) | 0/74 (0) | 2/96 (2.08) | 0/5 (0) |
| ST433-1LV | 0/24 (0) | 0/74 (0) | 0/96 (0) | 0/5 (0) |
| ST474 | 0/24 (0) | 0/74 (0) | 0/96 (0) | 0/5 (0) |
| ST48 | 0/24 (0) | 0/74 (0) | 0/96 (0) | 0/5 (0) |
| ST502 | 0/24 (0) | 1/74 (1.35) | 0/96 (0) | 0/5 (0) |
| ST557 | 0/24 (0) | 0/74 (0) | 0/96 (0) | 0/5 (0) |
| ST5785 | 0/24 (0) | 0/74 (0) | 1/96 (1.04) | 0/5 (0) |
| ST587 | 0/24 (0) | 0/74 (0) | 0/96 (0) | 0/5 (0) |
| ST607-1LV | 0/24 (0) | 0/74 (0) | 0/96 (0) | 0/5 (0) |
| ST628 | 0/24 (0) | 1/74 (1.35) | 0/96 (0) | 0/5 (0) |
| ST6580 | 0/24 (0) | 0/74 (0) | 0/96 (0) | 0/5 (0) |
| ST66 | 1/24 (4.17) | 0/74 (0) | 0/96 (0) | 0/5 (0) |
| ST664 | 0/24 (0) | 0/74 (0) | 0/96 (0) | 0/5 (0) |
| ST6792 | 0/24 (0) | 0/74 (0) | 0/96 (0) | 0/5 (0) |
| ST711 | 0/24 (0) | 1/74 (1.35) | 0/96 (0) | 0/5 (0) |
| ST872 | 0/24 (0) | 0/74 (0) | 0/96 (0) | 0/5 (0) |

| | | | | |
|---|---|---|---|---|
| **K-locus, n (%)** | | | | |
| KL1 | 0/24 (0) | 0/74 (0) | 0/96 (0) | 0/5 (0) |
| KL10 | 1/24 (4.17) | 0/74 (0) | 0/96 (0) | 0/5 (0) |
| KL104 | 0/24 (0) | 1/74 (1.35) | 0/96 (0) | 0/5 (0) |
| KL105 | 0/24 (0) | 2/74 (2.7) | 0/96 (0) | 0/5 (0) |
| KL106 | 0/24 (0) | 0/74 (0) | 0/96 (0) | 1/5 (20) |
| KL108 | 0/24 (0) | 0/74 (0) | 1/96 (1.04) | 0/5 (0) |
| KL109 | 0/24 (0) | 0/74 (0) | 0/96 (0) | 0/5 (0) |
| KL110 | 0/24 (0) | 0/74 (0) | 4/96 (4.17) | 0/5 (0) |
| KL112 | 0/24 (0) | 0/74 (0) | 1/96 (1.04) | 0/5 (0) |
| KL113 | 0/24 (0) | 1/74 (1.35) | 0/96 (0) | 0/5 (0) |
| KL114 | 0/24 (0) | 0/74 (0) | 0/96 (0) | 0/5 (0) |
| KL116 | 0/24 (0) | 1/74 (1.35) | 0/96 (0) | 0/5 (0) |
| KL122 | 0/24 (0) | 1/74 (1.35) | 0/96 (0) | 0/5 (0) |
| KL123 | 0/24 (0) | 1/74 (1.35) | 0/96 (0) | 0/5 (0) |
| KL132 | 0/24 (0) | 0/74 (0) | 0/96 (0) | 0/5 (0) |
| KL136 | 1/24 (4.17) | 2/74 (2.7) | 0/96 (0) | 0/5 (0) |
| KL140 | 0/24 (0) | 0/74 (0) | 0/96 (0) | 0/5 (0) |
| KL151 | 0/24 (0) | 0/74 (0) | 0/96 (0) | 0/5 (0) |
| KL16 | 0/24 (0) | 1/74 (1.35) | 1/96 (1.04) | 0/5 (0) |
| KL166 | 0/24 (0) | 0/74 (0) | 0/96 (0) | 0/5 (0) |
| KL17 | 0/24 (0) | 0/74 (0) | 0/96 (0) | 0/5 (0) |
| KL18 | 1/24 (4.17) | 0/74 (0) | 0/96 (0) | 0/5 (0) |
| KL19 | 0/24 (0) | 0/74 (0) | 0/96 (0) | 0/5 (0) |
| KL20 | 0/24 (0) | 0/74 (0) | 0/96 (0) | 0/5 (0) |
| KL22 | 0/24 (0) | 0/74 (0) | 0/96 (0) | 0/5 (0) |
| KL27 | 0/24 (0) | 0/74 (0) | 0/96 (0) | 0/5 (0) |
| KL28 | 0/24 (0) | 1/74 (1.35) | 2/96 (2.08) | 0/5 (0) |
| KL3 | 0/24 (0) | 1/74 (1.35) | 3/96 (3.12) | 0/5 (0) |
| KL38 | 2/24 (8.33) | 2/74 (2.7) | 0/96 (0) | 0/5 (0) |
| KL51 | 0/24 (0) | 0/74 (0) | 1/96 (1.04) | 0/5 (0) |
| KL52 | 0/24 (0) | 1/74 (1.35) | 0/96 (0) | 0/5 (0) |
| KL54 | 0/24 (0) | 2/74 (2.7) | 0/96 (0) | 0/5 (0) |
| KL55 | 0/24 (0) | 0/74 (0) | 0/96 (0) | 0/5 (0) |
| KL60 | 0/24 (0) | 0/74 (0) | 0/96 (0) | 0/5 (0) |
| KL63 | 0/24 (0) | 0/74 (0) | 0/96 (0) | 0/5 (0) |
| KL67 | 1/24 (4.17) | 1/74 (1.35) | 0/96 (0) | 0/5 (0) |
| KL7 | 0/24 (0) | 0/74 (0) | 1/96 (1.04) | 0/5 (0) |
| KL8 | 0/24 (0) | 0/74 (0) | 1/96 (1.04) | 0/5 (0) |
| KL9 | 0/24 (0) | 0/74 (0) | 0/96 (0) | 0/5 (0) |
| Unknown | 2/24 (8.33) | 3/74 (4.05) | 5/96 (5.21) | 0/5 (0) |
| **O-types, n (%)** | | | | |
| O13 | 1/24 (4.17) | 2/74 (2.7) | 0/96 (0) | 0/5 (0) |
| O1ab | 11/24 (45.83) | 33/74 (44.59) | 45/96 (46.88) | 5/5 (100) |
| O2a | 1/24 (4.17) | 1/74 (1.35) | 2/96 (2.08) | 0/5 (0) |
| O2afg | 4/24 (16.67) | 11/74 (14.86) | 23/96 (23.96) | 0/5 (0) |
| O3b | 0/24 (0) | 4/74 (5.41) | 4/96 (4.17) | 0/5 (0) |
| O4 | 0/24 (0) | 5/74 (6.76) | 5/96 (5.21) | 0/5 (0) |
| O5 | 7/24 (29.17) | 18/74 (24.32) | 17/96 (17.71) | 0/5 (0) |
| Unknown | 0/24 (0) | 0/74 (0) | 0/96 (0) | 0/5 (0) |

Data represent isolates collected from two hospital-based surveillance studies and two temporally aligned post-mortem studies.

**Supplementary Table 3: Clinical characteristics of infants up to 90 days of age with *K. pneumoniae* isolates that were sequenced**

| | All isolates (N=337) | pCAI (N=62) | pHAI (N=275) |
|---|---|---|---|
| **Median age in days (IQR)** | 12 (6-28) | 6.5 (2-25.75) | 12 (7-28.5) |
| **Type of infection, n (%)** | | | |
| Presumed hospital-associated | 62 (18.4) | 62 (100.0) | 0 (0.0) |
| Presumed community-acquired | 275 (81.6) | 0 (0.0) | 275 (100.0) |
| **Median number of days in hospital (IQR)** | 22 (6-55). N=293 | 5 (0-13.5). N=51 | 30.5 (10-62.75). N=242 |
| **Age group, n (%)** | | | |
| EOD | 14 (4.2) | 13 (21.0) | 1 (0.4) |
| LOD | 323 (95.8) | 49 (79.0) | 274 (99.6) |
| **Year of isolation, n (%)** | | | |
| 2018 | 24 (7.1) | 6 (9.7) | 18 (6.5) |
| 2019 | 66 (19.6) | 8 (12.9) | 58 (21.1) |
| 2020 | 66 (19.6) | 12 (19.4) | 54 (19.6) |
| 2021 | 27 (8.0) | 8 (12.9) | 19 (6.9) |
| 2022 | 54 (16.0) | 12 (19.4) | 42 (15.3) |
| 2023 | 100 (29.7) | 16 (25.8) | 84 (30.5) |
| **Gender, n (%)** | | | |
| Female | 144 (42.7) | 24 (38.7) | 120 (43.6) |
| Male | 191 (56.7) | 38 (61.3) | 153 (55.6) |
| Unknown | 2 (0.6) | 0 (0.0) | 2 (0.7) |
| **Mothers HIV status, n (%)** | | | |
| HIV uninfected | 226 (67.1) | 39 (62.9) | 187 (68.0) |
| HIV infected | 97 (28.8) | 15 (24.2) | 82 (29.8) |
| Unknown | 14 (4.2) | 8 (12.9) | 6 (2.2) |
| **Infant HIV status, n (%)** | | | |
| HIV uninfected | 196 (58.2) | 40 (64.5) | 156 (56.7) |
| HIV infected | 7 (2.1) | 2 (3.2) | 5 (1.8) |
| Unknown | 134 (39.8) | 20 (32.3) | 114 (41.5) |
| **Gestational age, n (%)** | | | |
| <28 weeks | 58 (17.2) | 4 (6.5) | 54 (19.6) |
| 28-<34 weeks | 147 (43.6) | 16 (25.8) | 131 (47.6) |
| 34-<37 weeks | 38 (11.3) | 7 (11.3) | 31 (11.3) |
| >=37 weeks | 47 (13.9) | 15 (24.2) | 32 (11.6) |
| Unknown | 47 (13.9) | 20 (32.3) | 27 (9.8) |
| **Birth weight, n (%)** | | | |
| <1000 grams | 77 (22.8) | 6 (9.7) | 71 (25.8) |
| 1000-<1500 grams | 94 (27.9) | 9 (14.5) | 85 (30.9) |
| 1500-<2500 grams | 84 (24.9) | 17 (27.4) | 67 (24.4) |
| 2500-<4000 grams | 49 (14.5) | 14 (22.6) | 35 (12.7) |
| >=4000 grams | 1 (0.3) | 1 (1.6) | 0 (0.0) |
| Unknown | 32 (9.5) | 15 (24.2) | 17 (6.2) |
| **Median birth weight in grams (IQR)** | 1355 (995-2055). N=305 | 2160 (1342.5-2735). N=47 | 1280 (985-1845). N=258 |
| **Antibiotics prelabour, n (%)** | 7 (2.1) | 1 (1.6) | 6 (2.2) |
| **Intrapartum antibiotics, n (%)** | 45 (13.4) | 2 (3.2) | 43 (15.6) |
| **Antibiotics during hospital admission, n (%)** | 122 (36.2) | 13 (21.0) | 109 (39.6) |

| | | | |
|---|---|---|---|
| **Infant outcome, n (%)** | | | |
| Discharged Home | 135 (40.1) | 22 (35.5) | 113 (41.1) |
| Died | 180 (53.4) | 36 (58.1) | 144 (52.4) |
| Unknown | 22 (6.5) | 4 (6.5) | 18 (6.5) |
| **Site of positive culture, n (%)** | | | |
| Blood | 236 (70.0) | 36 (58.1) | 200 (72.7) |
| CSF | 32 (9.5) | 12 (19.4) | 20 (7.3) |
| Lung tissue | 39 (11.6) | 10 (16.1) | 29 (10.5) |
| Unknown | 30 (8.9) | 4 (6.5) | 26 (9.5) |

Abbreviations: CSF, Cerebral spinal fluid; IQR, interquartile range; pCAI, presumed community acquires; pHAI, presumed hospital-associated

pCAI isolates were defined as invasive KPn detected on admission or within 72 hours of hospitalization or if the death occurred in the community.

pHAI was defined as an invasive KPn detected more than 72 hours after admission to the hospital or if the DeCoDe panel attributed nosocomial infection to the causal pathway of the death.

* Data only collected for postmortem studies.

IQR – interquartile range.

**Supplementary Table 4: Genomic characterization of *K. pneumoniae* causing invasive disease and mortality in South African infants 0-90 days of age, by collection site**

| | Bheki Mlangeni District Hospital (N=1) | Chris Hani Baragwanth Academic Hospital (N=247) | Charlotte Maxeke Johannesburg Academic Hospital (N=2) | Leratong hospital (N=7) | Mowbray Maternity Hospital (N=2) | Prince Mshiyeni Memorial Hospital (N=6) | Rahima Moosa Mother and Child Hospital (N=36) | Steve Biko Academic Hospital (N=11) | Thelle Mogoerane Regional Hospital (N=2) | Tshepo Temba Clinic (N=1) | Zola Clinic (N=1) | Unknown (N=2) |
|---|---|---|---|---|---|---|---|---|---|---|---|---|
| **Sequence type, n (%)** | | | | | | | | | | | | |
| ST14 | 0 (0) | 16 (6.48) | 0 (0) | 0 (0) | 0 (0) | 0 (0) | 2 (5.56) | 0 (0) | 0 (0) | 0 (0) | 0 (0) | 0 (0) |
| ST152 | 0 (0) | 7 (2.83) | 0 (0) | 0 (0) | 0 (0) | 0 (0) | 1 (2.78) | 3 (27.27) | 0 (0) | 0 (0) | 0 (0) | 0 (0) |
| ST17 | 0 (0) | 62 (25.1) | 1 (50) | 2 (28.57) | 0 (0) | 0 (0) | 5 (13.89) | 0 (0) | 0 (0) | 0 (0) | 1 (100) | 1 (50) |
| ST20 | 0 (0) | 2 (0.81) | 0 (0) | 2 (28.57) | 0 (0) | 0 (0) | 1 (2.78) | 0 (0) | 0 (0) | 0 (0) | 0 (0) | 0 (0) |
| ST25 | 0 (0) | 15 (6.07) | 0 (0) | 0 (0) | 0 (0) | 1 (16.67) | 1 (2.78) | 1 (9.09) | 0 (0) | 0 (0) | 0 (0) | 0 (0) |
| ST29 | 0 (0) | 5 (2.02) | 0 (0) | 2 (28.57) | 0 (0) | 0 (0) | 1 (2.78) | 0 (0) | 0 (0) | 0 (0) | 0 (0) | 0 (0) |
| ST307 | 0 (0) | 25 (10.12) | 0 (0) | 0 (0) | 0 (0) | 1 (16.67) | 0 (0) | 1 (9.09) | 0 (0) | 0 (0) | 0 (0) | 0 (0) |
| ST37 | 0 (0) | 7 (2.83) | 0 (0) | 0 (0) | 0 (0) | 0 (0) | 0 (0) | 0 (0) | 0 (0) | 0 (0) | 0 (0) | 0 (0) |
| ST39 | 0 (0) | 28 (11.34) | 0 (0) | 0 (0) | 1 (50) | 3 (50) | 4 (11.11) | 3 (27.27) | 0 (0) | 0 (0) | 0 (0) | 0 (0) |
| ST45 | 0 (0) | 4 (1.62) | 0 (0) | 0 (0) | 0 (0) | 0 (0) | 7 (19.44) | 0 (0) | 0 (0) | 0 (0) | 0 (0) | 0 (0) |
| ST607 | 0 (0) | 10 (4.05) | 0 (0) | 0 (0) | 0 (0) | 0 (0) | 0 (0) | 1 (9.09) | 0 (0) | 0 (0) | 0 (0) | 0 (0) |
| ST985 | 0 (0) | 8 (3.24) | 0 (0) | 0 (0) | 0 (0) | 0 (0) | 0 (0) | 0 (0) | 0 (0) | 0 (0) | 0 (0) | 0 (0) |
| ST101 | 0 (0) | 1 (0.4) | 0 (0) | 0 (0) | 0 (0) | 0 (0) | 0 (0) | 0 (0) | 2 (100) | 0 (0) | 0 (0) | 0 (0) |
| ST1207 | 0 (0) | 2 (0.81) | 0 (0) | 0 (0) | 0 (0) | 0 (0) | 0 (0) | 0 (0) | 0 (0) | 0 (0) | 0 (0) | 0 (0) |
| ST1296 | 0 (0) | 2 (0.81) | 0 (0) | 0 (0) | 0 (0) | 0 (0) | 0 (0) | 0 (0) | 0 (0) | 0 (0) | 0 (0) | 0 (0) |
| ST13 | 0 (0) | 4 (1.62) | 0 (0) | 0 (0) | 0 (0) | 0 (0) | 0 (0) | 0 (0) | 0 (0) | 0 (0) | 0 (0) | 0 (0) |
| ST133 | 0 (0) | 1 (0.4) | 0 (0) | 0 (0) | 0 (0) | 0 (0) | 0 (0) | 0 (0) | 0 (0) | 0 (0) | 0 (0) | 0 (0) |
| ST1380 | 0 (0) | 0 (0) | 0 (0) | 0 (0) | 0 (0) | 0 (0) | 1 (2.78) | 0 (0) | 0 (0) | 0 (0) | 0 (0) | 0 (0) |
| ST140 | 0 (0) | 0 (0) | 0 (0) | 0 (0) | 0 (0) | 0 (0) | 0 (0) | 0 (0) | 0 (0) | 1 (100) | 0 (0) | 0 (0) |
| ST1414 | 0 (0) | 1 (0.4) | 0 (0) | 0 (0) | 0 (0) | 1 (16.67) | 0 (0) | 0 (0) | 0 (0) | 0 (0) | 0 (0) | 0 (0) |
| ST1427 | 0 (0) | 2 (0.81) | 0 (0) | 0 (0) | 0 (0) | 0 (0) | 0 (0) | 0 (0) | 0 (0) | 0 (0) | 0 (0) | 0 (0) |
| ST1444 | 0 (0) | 2 (0.81) | 0 (0) | 0 (0) | 0 (0) | 0 (0) | 0 (0) | 0 (0) | 0 (0) | 0 (0) | 0 (0) | 0 (0) |
| ST15 | 0 (0) | 2 (0.81) | 0 (0) | 0 (0) | 0 (0) | 0 (0) | 0 (0) | 0 (0) | 0 (0) | 0 (0) | 0 (0) | 0 (0) |
| ST1552 | 0 (0) | 0 (0) | 0 (0) | 0 (0) | 0 (0) | 0 (0) | 2 (5.56) | 0 (0) | 0 (0) | 0 (0) | 0 (0) | 0 (0) |
| ST17-1LV | 0 (0) | 0 (0) | 0 (0) | 0 (0) | 0 (0) | 0 (0) | 0 (0) | 0 (0) | 0 (0) | 0 (0) | 0 (0) | 0 (0) |
| ST1873 | 0 (0) | 2 (0.81) | 0 (0) | 0 (0) | 0 (0) | 0 (0) | 0 (0) | 0 (0) | 0 (0) | 0 (0) | 0 (0) | 0 (0) |
| ST1898 | 0 (0) | 1 (0.4) | 0 (0) | 0 (0) | 0 (0) | 0 (0) | 0 (0) | 0 (0) | 0 (0) | 0 (0) | 0 (0) | 0 (0) |
| ST193 | 0 (0) | 0 (0) | 0 (0) | 0 (0) | 0 (0) | 0 (0) | 0 (0) | 1 (9.09) | 0 (0) | 0 (0) | 0 (0) | 0 (0) |
| ST2039 | 0 (0) | 1 (0.4) | 0 (0) | 0 (0) | 0 (0) | 0 (0) | 0 (0) | 0 (0) | 0 (0) | 0 (0) | 0 (0) | 0 (0) |
| ST219 | 0 (0) | 2 (0.81) | 0 (0) | 0 (0) | 0 (0) | 0 (0) | 0 (0) | 0 (0) | 0 (0) | 0 (0) | 0 (0) | 0 (0) |
| ST2191-1LV | 0 (0) | 1 (0.4) | 0 (0) | 0 (0) | 0 (0) | 0 (0) | 0 (0) | 0 (0) | 0 (0) | 0 (0) | 0 (0) | 0 (0) |
| ST22 | 0 (0) | 1 (0.4) | 0 (0) | 0 (0) | 0 (0) | 0 (0) | 0 (0) | 0 (0) | 0 (0) | 0 (0) | 0 (0) | 0 (0) |
| ST23 | 0 (0) | 0 (0) | 0 (0) | 0 (0) | 0 (0) | 0 (0) | 0 (0) | 0 (0) | 0 (0) | 0 (0) | 0 (0) | 0 (0) |
| ST2441 | 0 (0) | 1 (0.4) | 0 (0) | 0 (0) | 0 (0) | 0 (0) | 0 (0) | 0 (0) | 0 (0) | 0 (0) | 0 (0) | 0 (0) |
| ST25-1LV | 0 (0) | 3 (1.21) | 0 (0) | 0 (0) | 0 (0) | 0 (0) | 0 (0) | 0 (0) | 0 (0) | 0 (0) | 0 (0) | 0 (0) |
| ST252 | 0 (0) | 0 (0) | 0 (0) | 0 (0) | 0 (0) | 0 (0) | 0 (0) | 1 (9.09) | 0 (0) | 0 (0) | 0 (0) | 0 (0) |
| ST253 | 0 (0) | 2 (0.81) | 0 (0) | 0 (0) | 0 (0) | 0 (0) | 0 (0) | 0 (0) | 0 (0) | 0 (0) | 0 (0) | 0 (0) |
| ST26 | 0 (0) | 0 (0) | 0 (0) | 0 (0) | 0 (0) | 0 (0) | 0 (0) | 0 (0) | 0 (0) | 0 (0) | 0 (0) | 0 (0) |

| | | | | | | | | | | | | |
|---|---|---|---|---|---|---|---|---|---|---|---|---|
| ST268 | 0 (0) | 1 (0.4) | 0 (0) | 0 (0) | 0 (0) | 0 (0) | 0 (0) | 0 (0) | 0 (0) | 0 (0) | 0 (0) | 0 (0) |
| ST294-2LV | 0 (0) | 0 (0) | 0 (0) | 0 (0) | 0 (0) | 0 (0) | 1 (2.78) | 0 (0) | 0 (0) | 0 (0) | 0 (0) | 0 (0) |
| ST297-1LV | 0 (0) | 1 (0.4) | 0 (0) | 0 (0) | 0 (0) | 0 (0) | 0 (0) | 0 (0) | 0 (0) | 0 (0) | 0 (0) | 0 (0) |
| ST3073 | 0 (0) | 1 (0.4) | 0 (0) | 0 (0) | 0 (0) | 0 (0) | 0 (0) | 0 (0) | 0 (0) | 0 (0) | 0 (0) | 0 (0) |
| ST321 | 0 (0) | 0 (0) | 1 (50) | 0 (0) | 0 (0) | 0 (0) | 0 (0) | 0 (0) | 0 (0) | 0 (0) | 0 (0) | 0 (0) |
| ST3280 | 0 (0) | 1 (0.4) | 0 (0) | 0 (0) | 0 (0) | 0 (0) | 0 (0) | 0 (0) | 0 (0) | 0 (0) | 0 (0) | 0 (0) |
| ST336 | 0 (0) | 0 (0) | 0 (0) | 0 (0) | 0 (0) | 0 (0) | 1 (2.78) | 0 (0) | 0 (0) | 0 (0) | 0 (0) | 0 (0) |
| ST34 | 0 (0) | 0 (0) | 0 (0) | 0 (0) | 0 (0) | 0 (0) | 1 (2.78) | 0 (0) | 0 (0) | 0 (0) | 0 (0) | 0 (0) |
| ST348 | 0 (0) | 0 (0) | 0 (0) | 0 (0) | 0 (0) | 0 (0) | 1 (2.78) | 0 (0) | 0 (0) | 0 (0) | 0 (0) | 0 (0) |
| ST3483-1LV | 0 (0) | 1 (0.4) | 0 (0) | 0 (0) | 0 (0) | 0 (0) | 0 (0) | 0 (0) | 0 (0) | 0 (0) | 0 (0) | 0 (0) |
| ST35 | 0 (0) | 2 (0.81) | 0 (0) | 0 (0) | 0 (0) | 0 (0) | 1 (2.78) | 0 (0) | 0 (0) | 0 (0) | 0 (0) | 0 (0) |
| ST353 | 0 (0) | 0 (0) | 0 (0) | 0 (0) | 0 (0) | 0 (0) | 4 (11.11) | 0 (0) | 0 (0) | 0 (0) | 0 (0) | 0 (0) |
| ST380 | 0 (0) | 1 (0.4) | 0 (0) | 0 (0) | 0 (0) | 0 (0) | 0 (0) | 0 (0) | 0 (0) | 0 (0) | 0 (0) | 0 (0) |
| ST399 | 0 (0) | 1 (0.4) | 0 (0) | 0 (0) | 0 (0) | 0 (0) | 0 (0) | 0 (0) | 0 (0) | 0 (0) | 0 (0) | 0 (0) |
| ST405 | 0 (0) | 1 (0.4) | 0 (0) | 0 (0) | 0 (0) | 0 (0) | 0 (0) | 0 (0) | 0 (0) | 0 (0) | 0 (0) | 0 (0) |
| ST416 | 1 (100) | 0 (0) | 0 (0) | 0 (0) | 0 (0) | 0 (0) | 0 (0) | 0 (0) | 0 (0) | 0 (0) | 0 (0) | 0 (0) |
| ST4291 | 0 (0) | 2 (0.81) | 0 (0) | 0 (0) | 0 (0) | 0 (0) | 0 (0) | 0 (0) | 0 (0) | 0 (0) | 0 (0) | 0 (0) |
| ST433-1LV | 0 (0) | 1 (0.4) | 0 (0) | 0 (0) | 0 (0) | 0 (0) | 0 (0) | 0 (0) | 0 (0) | 0 (0) | 0 (0) | 0 (0) |
| ST474 | 0 (0) | 1 (0.4) | 0 (0) | 0 (0) | 0 (0) | 0 (0) | 0 (0) | 0 (0) | 0 (0) | 0 (0) | 0 (0) | 0 (0) |
| ST48 | 0 (0) | 1 (0.4) | 0 (0) | 0 (0) | 0 (0) | 0 (0) | 0 (0) | 0 (0) | 0 (0) | 0 (0) | 0 (0) | 0 (0) |
| ST502 | 0 (0) | 5 (2.02) | 0 (0) | 1 (14.29) | 0 (0) | 0 (0) | 0 (0) | 0 (0) | 0 (0) | 0 (0) | 0 (0) | 0 (0) |
| ST557 | 0 (0) | 1 (0.4) | 0 (0) | 0 (0) | 0 (0) | 0 (0) | 0 (0) | 0 (0) | 0 (0) | 0 (0) | 0 (0) | 0 (0) |
| ST5785 | 0 (0) | 1 (0.4) | 0 (0) | 0 (0) | 0 (0) | 0 (0) | 0 (0) | 0 (0) | 0 (0) | 0 (0) | 0 (0) | 0 (0) |
| ST587 | 0 (0) | 1 (0.4) | 0 (0) | 0 (0) | 0 (0) | 0 (0) | 0 (0) | 0 (0) | 0 (0) | 0 (0) | 0 (0) | 0 (0) |
| ST607-1LV | 0 (0) | 1 (0.4) | 0 (0) | 0 (0) | 0 (0) | 0 (0) | 0 (0) | 0 (0) | 0 (0) | 0 (0) | 0 (0) | 0 (0) |
| ST628 | 0 (0) | 0 (0) | 0 (0) | 0 (0) | 0 (0) | 0 (0) | 1 (2.78) | 0 (0) | 0 (0) | 0 (0) | 0 (0) | 1 (50) |
| ST6580 | 0 (0) | 1 (0.4) | 0 (0) | 0 (0) | 0 (0) | 0 (0) | 0 (0) | 0 (0) | 0 (0) | 0 (0) | 0 (0) | 0 (0) |
| ST66 | 0 (0) | 0 (0) | 0 (0) | 0 (0) | 0 (0) | 0 (0) | 0 (0) | 0 (0) | 0 (0) | 0 (0) | 0 (0) | 0 (0) |
| ST664 | 0 (0) | 1 (0.4) | 0 (0) | 0 (0) | 0 (0) | 0 (0) | 0 (0) | 0 (0) | 0 (0) | 0 (0) | 0 (0) | 0 (0) |
| ST6792 | 0 (0) | 1 (0.4) | 0 (0) | 0 (0) | 0 (0) | 0 (0) | 0 (0) | 0 (0) | 0 (0) | 0 (0) | 0 (0) | 0 (0) |
| ST711 | 0 (0) | 0 (0) | 0 (0) | 0 (0) | 1 (50) | 0 (0) | 0 (0) | 0 (0) | 0 (0) | 0 (0) | 0 (0) | 0 (0) |
| ST872 | 0 (0) | 0 (0) | 0 (0) | 0 (0) | 0 (0) | 0 (0) | 1 (2.78) | 0 (0) | 0 (0) | 0 (0) | 0 (0) | 0 (0) |
| K-loci, n (%) | | | | | | | | | | | | |
| KL102 | 0 (0) | 28 (11.34) | 0 (0) | 0 (0) | 0 (0) | 1 (16.67) | 0 (0) | 1 (9.09) | 0 (0) | 0 (0) | 0 (0) | 0 (0) |
| KL149 | 0 (0) | 33 (13.36) | 0 (0) | 0 (0) | 0 (0) | 3 (50) | 3 (8.33) | 3 (27.27) | 0 (0) | 0 (0) | 0 (0) | 0 (0) |
| KL15 | 0 (0) | 12 (4.86) | 0 (0) | 1 (14.29) | 0 (0) | 0 (0) | 0 (0) | 0 (0) | 0 (0) | 0 (0) | 0 (0) | 0 (0) |
| KL2 | 0 (0) | 37 (14.98) | 0 (0) | 0 (0) | 1 (50) | 1 (16.67) | 5 (13.89) | 3 (27.27) | 0 (0) | 0 (0) | 0 (0) | 0 (0) |
| KL24 | 0 (0) | 4 (1.62) | 0 (0) | 0 (0) | 0 (0) | 0 (0) | 7 (19.44) | 0 (0) | 0 (0) | 0 (0) | 0 (0) | 0 (0) |
| KL25 | 0 (0) | 66 (26.72) | 1 (50) | 2 (28.57) | 0 (0) | 0 (0) | 7 (19.44) | 1 (9.09) | 0 (0) | 0 (0) | 0 (0) | 1 (50) |
| KL30 | 1 (100) | 3 (1.21) | 0 (0) | 2 (28.57) | 0 (0) | 0 (0) | 1 (2.78) | 1 (9.09) | 0 (0) | 0 (0) | 0 (0) | 0 (0) |
| KL39 | 0 (0) | 8 (3.24) | 0 (0) | 0 (0) | 0 (0) | 0 (0) | 0 (0) | 0 (0) | 0 (0) | 0 (0) | 0 (0) | 0 (0) |
| KL62 | 0 (0) | 8 (3.24) | 0 (0) | 0 (0) | 0 (0) | 0 (0) | 1 (2.78) | 1 (9.09) | 0 (0) | 0 (0) | 1 (100) | 0 (0) |
| KL1 | 0 (0) | 0 (0) | 0 (0) | 0 (0) | 0 (0) | 0 (0) | 0 (0) | 0 (0) | 0 (0) | 0 (0) | 0 (0) | 0 (0) |
| KL10 | 0 (0) | 3 (1.21) | 0 (0) | 0 (0) | 0 (0) | 0 (0) | 0 (0) | 0 (0) | 0 (0) | 0 (0) | 0 (0) | 0 (0) |
| KL104 | 0 (0) | 0 (0) | 0 (0) | 0 (0) | 0 (0) | 0 (0) | 1 (2.78) | 0 (0) | 0 (0) | 0 (0) | 0 (0) | 0 (0) |
| KL105 | 0 (0) | 2 (0.81) | 0 (0) | 0 (0) | 0 (0) | 0 (0) | 0 (0) | 0 (0) | 0 (0) | 0 (0) | 0 (0) | 0 (0) |
| KL106 | 0 (0) | 1 (0.4) | 0 (0) | 0 (0) | 0 (0) | 0 (0) | 0 (0) | 0 (0) | 0 (0) | 0 (0) | 0 (0) | 0 (0) |
| KL108 | 0 (0) | 1 (0.4) | 0 (0) | 0 (0) | 0 (0) | 0 (0) | 0 (0) | 0 (0) | 0 (0) | 0 (0) | 0 (0) | 0 (0) |

| | | | | | | | | | | | | |
|---|---|---|---|---|---|---|---|---|---|---|---|---|
| KL109 | 0 (0) | 0 (0) | 0 (0) | 0 (0) | 0 (0) | 0 (0) | 1 (2.78) | 0 (0) | 0 (0) | 0 (0) | 0 (0) | 0 (0) |
| KL110 | 0 (0) | 0 (0) | 0 (0) | 0 (0) | 0 (0) | 0 (0) | 4 (11.11) | 0 (0) | 0 (0) | 0 (0) | 0 (0) | 0 (0) |
| KL112 | 0 (0) | 1 (0.4) | 0 (0) | 0 (0) | 0 (0) | 0 (0) | 0 (0) | 0 (0) | 0 (0) | 0 (0) | 0 (0) | 0 (0) |
| KL113 | 0 (0) | 1 (0.4) | 0 (0) | 0 (0) | 0 (0) | 0 (0) | 0 (0) | 0 (0) | 0 (0) | 0 (0) | 0 (0) | 0 (0) |
| KL114 | 0 (0) | 2 (0.81) | 0 (0) | 0 (0) | 0 (0) | 0 (0) | 0 (0) | 0 (0) | 0 (0) | 0 (0) | 0 (0) | 0 (0) |
| KL116 | 0 (0) | 2 (0.81) | 0 (0) | 0 (0) | 0 (0) | 0 (0) | 0 (0) | 0 (0) | 0 (0) | 0 (0) | 0 (0) | 0 (0) |
| KL122 | 0 (0) | 0 (0) | 0 (0) | 0 (0) | 0 (0) | 0 (0) | 1 (2.78) | 0 (0) | 0 (0) | 0 (0) | 0 (0) | 0 (0) |
| KL123 | 0 (0) | 0 (0) | 0 (0) | 0 (0) | 0 (0) | 0 (0) | 1 (2.78) | 0 (0) | 0 (0) | 0 (0) | 0 (0) | 1 (50) |
| KL132 | 0 (0) | 1 (0.4) | 0 (0) | 0 (0) | 0 (0) | 0 (0) | 0 (0) | 0 (0) | 0 (0) | 0 (0) | 0 (0) | 0 (0) |
| KL136 | 0 (0) | 3 (1.21) | 0 (0) | 0 (0) | 0 (0) | 0 (0) | 1 (2.78) | 0 (0) | 0 (0) | 0 (0) | 0 (0) | 0 (0) |
| KL140 | 0 (0) | 1 (0.4) | 0 (0) | 0 (0) | 0 (0) | 0 (0) | 0 (0) | 0 (0) | 0 (0) | 0 (0) | 0 (0) | 0 (0) |
| KL151 | 0 (0) | 1 (0.4) | 0 (0) | 0 (0) | 0 (0) | 0 (0) | 0 (0) | 0 (0) | 0 (0) | 0 (0) | 0 (0) | 0 (0) |
| KL16 | 0 (0) | 1 (0.4) | 0 (0) | 0 (0) | 0 (0) | 0 (0) | 1 (2.78) | 0 (0) | 0 (0) | 0 (0) | 0 (0) | 0 (0) |
| KL166 | 0 (0) | 1 (0.4) | 0 (0) | 0 (0) | 0 (0) | 0 (0) | 0 (0) | 0 (0) | 0 (0) | 0 (0) | 0 (0) | 0 (0) |
| KL17 | 0 (0) | 1 (0.4) | 0 (0) | 0 (0) | 0 (0) | 0 (0) | 0 (0) | 0 (0) | 2 (100) | 0 (0) | 0 (0) | 0 (0) |
| KL18 | 0 (0) | 1 (0.4) | 0 (0) | 0 (0) | 0 (0) | 0 (0) | 0 (0) | 0 (0) | 0 (0) | 0 (0) | 0 (0) | 0 (0) |
| KL19 | 0 (0) | 3 (1.21) | 0 (0) | 0 (0) | 0 (0) | 0 (0) | 0 (0) | 0 (0) | 0 (0) | 0 (0) | 0 (0) | 0 (0) |
| KL20 | 0 (0) | 1 (0.4) | 0 (0) | 0 (0) | 0 (0) | 0 (0) | 0 (0) | 0 (0) | 0 (0) | 0 (0) | 0 (0) | 0 (0) |
| KL22 | 0 (0) | 1 (0.4) | 0 (0) | 0 (0) | 0 (0) | 0 (0) | 0 (0) | 0 (0) | 0 (0) | 0 (0) | 0 (0) | 0 (0) |
| KL27 | 0 (0) | 1 (0.4) | 0 (0) | 0 (0) | 0 (0) | 0 (0) | 0 (0) | 0 (0) | 0 (0) | 0 (0) | 0 (0) | 0 (0) |
| KL28 | 0 (0) | 3 (1.21) | 0 (0) | 2 (28.57) | 0 (0) | 0 (0) | 1 (2.78) | 0 (0) | 0 (0) | 0 (0) | 0 (0) | 0 (0) |
| KL3 | 0 (0) | 4 (1.62) | 1 (50) | 0 (0) | 0 (0) | 0 (0) | 0 (0) | 0 (0) | 0 (0) | 0 (0) | 0 (0) | 0 (0) |
| KL38 | 0 (0) | 2 (0.81) | 0 (0) | 0 (0) | 0 (0) | 0 (0) | 0 (0) | 0 (0) | 0 (0) | 0 (0) | 0 (0) | 0 (0) |
| KL51 | 0 (0) | 0 (0) | 0 (0) | 0 (0) | 0 (0) | 0 (0) | 0 (0) | 1 (9.09) | 0 (0) | 0 (0) | 0 (0) | 0 (0) |
| KL52 | 0 (0) | 1 (0.4) | 0 (0) | 0 (0) | 0 (0) | 0 (0) | 0 (0) | 0 (0) | 0 (0) | 0 (0) | 0 (0) | 0 (0) |
| KL54 | 0 (0) | 1 (0.4) | 0 (0) | 0 (0) | 1 (50) | 0 (0) | 0 (0) | 0 (0) | 0 (0) | 0 (0) | 0 (0) | 0 (0) |
| KL55 | 0 (0) | 2 (0.81) | 0 (0) | 0 (0) | 0 (0) | 0 (0) | 0 (0) | 0 (0) | 0 (0) | 1 (100) | 0 (0) | 0 (0) |
| KL60 | 0 (0) | 1 (0.4) | 0 (0) | 0 (0) | 0 (0) | 0 (0) | 0 (0) | 0 (0) | 0 (0) | 0 (0) | 0 (0) | 0 (0) |
| KL63 | 0 (0) | 0 (0) | 0 (0) | 0 (0) | 0 (0) | 0 (0) | 1 (2.78) | 0 (0) | 0 (0) | 0 (0) | 0 (0) | 0 (0) |
| KL67 | 0 (0) | 2 (0.81) | 0 (0) | 0 (0) | 0 (0) | 0 (0) | 0 (0) | 0 (0) | 0 (0) | 0 (0) | 0 (0) | 0 (0) |
| KL7 | 0 (0) | 1 (0.4) | 0 (0) | 0 (0) | 0 (0) | 0 (0) | 0 (0) | 0 (0) | 0 (0) | 0 (0) | 0 (0) | 0 (0) |
| KL8 | 0 (0) | 1 (0.4) | 0 (0) | 0 (0) | 0 (0) | 1 (16.67) | 0 (0) | 0 (0) | 0 (0) | 0 (0) | 0 (0) | 0 (0) |
| KL9 | 0 (0) | 1 (0.4) | 0 (0) | 0 (0) | 0 (0) | 0 (0) | 0 (0) | 0 (0) | 0 (0) | 0 (0) | 0 (0) | 0 (0) |
| Unknown | 0 (0) | 7 (2.83) | 1 (50) | 0 (0) | 0 (0) | 0 (0) | 1 (2.78) | 1 (9.09) | 0 (0) | 1 (100) | 0 (0) | 0 (0) |
| O-types, n (%) | | | | | | | | | | | | |
| O13 | 0 (0) | 3 (1.21) | 0 (0) | 0 (0) | 0 (0) | 0 (0) | 0 (0) | 0 (0) | 0 (0) | 0 (0) | 0 (0) | 0 (0) |
| O1ab | 1 (100) | 115 (46.56) | 0 (0) | 4 (57.14) | 2 (100) | 4 (66.67) | 16 (44.44) | 7 (63.64) | 2 (100) | 1 (100) | 0 (0) | 0 (0) |
| O2a | 0 (0) | 7 (2.83) | 0 (0) | 0 (0) | 0 (0) | 0 (0) | 7 (19.44) | 0 (0) | 0 (0) | 0 (0) | 0 (0) | 0 (0) |
| O2afg | 0 (0) | 42 (17) | 1 (50) | 0 (0) | 0 (0) | 2 (33.33) | 2 (5.56) | 1 (9.09) | 0 (0) | 0 (0) | 1 (100) | 0 (0) |
| O3b | 0 (0) | 4 (1.62) | 0 (0) | 0 (0) | 0 (0) | 0 (0) | 5 (13.89) | 0 (0) | 0 (0) | 0 (0) | 0 (0) | 1 (50) |
| O4 | 0 (0) | 21 (8.5) | 0 (0) | 1 (14.29) | 0 (0) | 0 (0) | 1 (2.78) | 3 (27.27) | 0 (0) | 0 (0) | 0 (0) | 0 (0) |
| O5 | 0 (0) | 55 (22.27) | 1 (50) | 2 (28.57) | 0 (0) | 0 (0) | 5 (13.89) | 0 (0) | 0 (0) | 0 (0) | 0 (0) | 1 (50) |
| Unknown | 0 (0) | 0 (0) | 0 (0) | 0 (0) | 0 (0) | 0 (0) | 0 (0) | 0 (0) | 0 (0) | 0 (0) | 0 (0) | 0 (0) |
| Multi-drug resistance (MDR), n (%) | 0 (0) | 214 (86.64) | 1 (50) | 6 (85.71) | 2 (100) | 6 (100) | 29 (80.56) | 11 (100) | 2 (100) | 0 (0) | 1 (100) | 1 (50) |
| Aminoglycosides | 1 (100) | 211 (85.43) | 1 (50) | 6 (85.71) | 2 (100) | 6 (100) | 30 (83.33) | 10 (90.91) | 2 (100) | 0 (0) | 1 (100) | 1 (50) |
| Carbapenems | 0 (0) | 83 (33.6) | 1 (50) | 7 (100) | 0 (0) | 3 (50) | 6 (16.67) | 7 (63.64) | 0 (0) | 0 (0) | 0 (0) | 0 (0) |
| 3rd Generation Cephalosporins | 0 (0) | 196 (79.35) | 1 (50) | 6 (85.71) | 2 (100) | 6 (100) | 20 (55.56) | 9 (81.82) | 2 (100) | 0 (0) | 1 (100) | 1 (50) |
| Colistin | 0 (0) | 4 (1.62) | 0 (0) | 1 (14.29) | 0 (0) | 0 (0) | 2 (5.56) | 0 (0) | 0 (0) | 0 (0) | 0 (0) | 0 (0) |

| | | | | | | | | | | | | |
|---|---|---|---|---|---|---|---|---|---|---|---|---|
| Fluoroquinolones | 0 (0) | 118 (47.77) | 1 (50) | 6 (85.71) | 0 (0) | 5 (83.33) | 18 (50) | 9 (81.82) | 2 (100) | 0 (0) | 0 (0) | 0 (0) |
| Fosfomycin | 0 (0) | 0 (0) | 0 (0) | 0 (0) | 0 (0) | 0 (0) | 0 (0) | 0 (0) | 0 (0) | 0 (0) | 0 (0) | 0 (0) |
| Penicillins | 0 (0) | 240 (97.17) | 2 (100) | 7 (100) | 2 (100) | 5 (83.33) | 36 (100) | 11 (100) | 2 (100) | 1 (100) | 1 (100) | 2 (100) |
| Penicillin (beta-lactamase inhibitors) | 0 (0) | 2 (0.81) | 0 (0) | 0 (0) | 0 (0) | 0 (0) | 0 (0) | 0 (0) | 0 (0) | 0 (0) | 0 (0) | 0 (0) |
| Amphenicols | 0 (0) | 160 (64.78) | 1 (50) | 1 (14.29) | 2 (100) | 4 (66.67) | 18 (50) | 6 (54.55) | 0 (0) | 0 (0) | 0 (0) | 1 (50) |
| Sulfonamides | 0 (0) | 183 (74.09) | 1 (50) | 6 (85.71) | 2 (100) | 6 (100) | 28 (77.78) | 9 (81.82) | 2 (100) | 0 (0) | 0 (0) | 1 (50) |
| Tetracycline | 0 (0) | 47 (19.03) | 0 (0) | 2 (28.57) | 1 (50) | 1 (16.67) | 11 (30.56) | 4 (36.36) | 2 (100) | 0 (0) | 0 (0) | 0 (0) |
| Tigecycline | 0 (0) | 0 (0) | 0 (0) | 0 (0) | 0 (0) | 0 (0) | 0 (0) | 0 (0) | 0 (0) | 0 (0) | 0 (0) | 0 (0) |
| Trimethoprim | 0 (0) | 185 (74.9) | 1 (50) | 6 (85.71) | 2 (100) | 6 (100) | 28 (77.78) | 8 (72.73) | 2 (100) | 0 (0) | 0 (0) | 1 (50) |
| Virulence factors, n (%) | | | | | | | | | | | | |
| ybt | 0 (0) | 167 (67.61) | 1 (50) | 2 (28.57) | 1 (50) | 4 (66.67) | 24 (66.67) | 6 (54.55) | 0 (0) | 0 (0) | 0 (0) | 1 (50) |
| clb | 0 (0) | 1 (0.4) | 0 (0) | 0 (0) | 1 (50) | 0 (0) | 0 (0) | 0 (0) | 0 (0) | 0 (0) | 0 (0) | 0 (0) |
| iuc | 0 (0) | 2 (0.81) | 0 (0) | 0 (0) | 0 (0) | 0 (0) | 0 (0) | 0 (0) | 0 (0) | 0 (0) | 0 (0) | 0 (0) |
| Virulence factor combinations, n (%) | | | | | | | | | | | | |
| ybt only | 0 (0) | 165 (66.8) | 1 (50) | 2 (28.57) | 1 (50) | 4 (66.67) | 24 (66.67) | 6 (54.55) | 0 (0) | 0 (0) | 0 (0) | 1 (50) |
| ybt and clb | 0 (0) | 1 (0.4) | 0 (0) | 0 (0) | 1 (50) | 0 (0) | 0 (0) | 0 (0) | 0 (0) | 0 (0) | 0 (0) | 0 (0) |
| iuc only | 0 (0) | 0 (0) | 0 (0) | 0 (0) | 0 (0) | 0 (0) | 0 (0) | 0 (0) | 0 (0) | 0 (0) | 0 (0) | 0 (0) |
| icu and ybt w/o clb | 0 (0) | 1 (0.4) | 0 (0) | 0 (0) | 0 (0) | 0 (0) | 0 (0) | 0 (0) | 0 (0) | 0 (0) | 0 (0) | 0 (0) |
| ybt, clb and iuc | 0 (0) | 1 (0.4) | 0 (0) | 0 (0) | 0 (0) | 0 (0) | 0 (0) | 0 (0) | 0 (0) | 0 (0) | 0 (0) | 0 (0) |
| AMR and virulence factor combinations, n (%) | | | | | | | | | | | | |
| Carbapenems and ybt | 0 (0) | 60 (24.29) | 1 (50) | 2 (28.57) | 0 (0) | 3 (50) | 4 (11.11) | 4 (36.36) | 0 (0) | 0 (0) | 0 (0) | 0 (0) |
| Carbapenems and clb | 0 (0) | 0 (0) | 0 (0) | 0 (0) | 0 (0) | 0 (0) | 0 (0) | 0 (0) | 0 (0) | 0 (0) | 0 (0) | 0 (0) |
| ESBL and ybt | 0 (0) | 144 (58.3) | 1 (50) | 2 (28.57) | 1 (50) | 4 (66.67) | 16 (44.44) | 6 (54.55) | 0 (0) | 0 (0) | 0 (0) | 1 (50) |
| ESBL and clb | 0 (0) | 0 (0) | 0 (0) | 0 (0) | 1 (50) | 0 (0) | 0 (0) | 0 (0) | 0 (0) | 0 (0) | 0 (0) | 0 (0) |

Abbreviations: clb = Colibactin, ESBL = Extended Spectrum β-lactamase, iuc = Aerobactin, ST = Sequence type, ybt = Yersiniabactin.

#### Supplementary Table 5: Antimicrobial resistance genes

| ARGs | Overall (n=337) | pCAI (n=62) | pHAI (n=275) | OR (95% CI); p-value | aOR (95% CI); p-value |
|---|---|---|---|---|---|
| CMY-4.v1 | 1 (0.3) | 1 (1.61) | 0 (0) | 0.00 (0.00-8.79); 0.184 | - |
| OXA-1 | 149 (44.21) | 22 (35.48) | 127 (46.18) | 1.56 (0.85-2.91); 0.157 | 1.86 (0.99-3.64); 0.06 |
| DHA-1 | 9 (2.67) | 4 (6.45) | 5 (1.82) | 0.27 (0.06-1.40); 0.063 | - |
| OXA-10 | 3 (0.89) | 0 (0) | 3 (1.09) | - | - |
| TEM-1D.v1^ | 207 (61.42) | 30 (48.39) | 177 (64.36) | 1.92 (1.06-3.49); 0.021 | 2.97 (1.57-5.75); NA |
| SCO-1 | 12 (3.56) | 2 (3.23) | 10 (3.64) | 1.13 (0.23-10.89); >0.999 | - |
| OXA-1* | 2 (0.59) | 1 (1.61) | 1 (0.36) | 0.22 (0.00-17.76); 0.335 | - |
| OXA-9.v1 | 2 (0.59) | 0 (0) | 2 (0.73) | - | - |
| TEM-33 | 1 (0.3) | 1 (1.61) | 0 (0) | 0.00 (0.00-8.79); 0.184 | - |
| TEM-34 | 1 (0.3) | 0 (0) | 1 (0.36) | - | - |
| CTX-M-15 | 253 (75.07) | 39 (62.9) | 214 (77.82) | 2.06 (1.09-3.86); 0.022 | 2.99 (1.56-5.73); NA |
| CTX-M-15* | 1 (0.3) | 1 (1.61) | 0 (0) | 0.00 (0.00-8.79); 0.184 | - |
| CTX-M-3 | 2 (0.59) | 0 (0) | 2 (0.73) | - | - |
| SHV-12* | 1 (0.3) | 0 (0) | 1 (0.36) | - | - |
| NDM-1 | 7 (2.08) | 3 (4.84) | 4 (1.45) | 0.29 (0.05-2.04); 0.119 | 0.69 (0.11-6.85); 0.709 |
| OXA-181 | 89 (26.41) | 13 (20.97) | 76 (27.64) | 1.44 (0.72-3.06); 0.34 | 1.87 (0.86-4.51); 0.133 |
| NDM-5 | 1 (0.3) | 0 (0) | 1 (0.36) | - | - |
| VIM-1 | 8 (2.37) | 2 (3.23) | 6 (2.18) | 0.67 (0.12-6.95); 0.643 | - |
| OXA-23 | 1 (0.3) | 0 (0) | 1 (0.36) | - | - |
| OXA-66 | 1 (0.3) | 0 (0) | 1 (0.36) | - | - |
| OXA-48 | 1 (0.3) | 0 (0) | 1 (0.36) | - | - |
| SHV-1 | 10 (2.97) | 4 (6.45) | 6 (2.18) | 0.32 (0.07-1.62); 0.091 | - |
| SHV-1^ | 50 (14.84) | 14 (22.58) | 36 (13.09) | 0.52 (0.25-1.12); 0.074 | 0.68 (0.32-1.57); 0.342 |
| SHV-101* | 2 (0.59) | 0 (0) | 2 (0.73) | - | - |
| SHV-107* | 1 (0.3) | 1 (1.61) | 0 (0) | 0.00 (0.00-8.79); 0.184 | - |
| SHV-11.v1^ | 1 (0.3) | 0 (0) | 1 (0.36) | - | - |
| SHV-11^ | 165 (48.96) | 25 (40.32) | 140 (50.91) | 1.53 (0.85-2.81); 0.16 | 1.24 (0.66-2.34); 0.505 |
| SHV-13* | 1 (0.3) | 0 (0) | 1 (0.36) | - | - |
| SHV-172*? | 1 (0.3) | 1 (1.61) | 0 (0) | 0.00 (0.00-8.79); 0.184 | - |
| SHV-187 | 13 (3.86) | 2 (3.23) | 11 (4) | 1.25 (0.26-11.89); >0.999 | - |
| SHV-187* | 5 (1.48) | 1 (1.61) | 4 (1.45) | 0.90 (0.09-45.06); >0.999 | - |
| SHV-187^ | 1 (0.3) | 0 (0) | 1 (0.36) | - | - |
| SHV-207 | 1 (0.3) | 1 (1.61) | 0 (0) | 0.00 (0.00-8.79); 0.184 | - |
| SHV-209* | 1 (0.3) | 1 (1.61) | 0 (0) | 0.00 (0.00-8.79); 0.184 | - |
| SHV-220 | 1 (0.3) | 0 (0) | 1 (0.36) | - | - |
| SHV-26 | 4 (1.19) | 1 (1.61) | 3 (1.09) | 0.67 (0.05-35.89); 0.558 | - |
| SHV-27 | 1 (0.3) | 0 (0) | 1 (0.36) | - | - |
| SHV-28^ | 49 (14.54) | 5 (8.06) | 44 (16) | 2.17 (0.81-7.32); 0.161 | 5.15 (1.42-34.16); 0.035 |
| SHV-32 | 2 (0.59) | 0 (0) | 2 (0.73) | - | - |
| SHV-33 | 3 (0.89) | 2 (3.23) | 1 (0.36) | 0.11 (0.00-2.16); 0.088 | - |
| SHV-33* | 1 (0.3) | 0 (0) | 1 (0.36) | - | - |
| SHV-36 | 3 (0.89) | 0 (0) | 3 (1.09) | - | - |

| | | | | | |
|---|---|---|---|---|---|
| SHV-61 | 1 (0.3) | 0 (0) | 1 (0.36) | - | - |
| SHV-62 | 6 (1.78) | 2 (3.23) | 4 (1.45) | 0.44 (0.06-5.02); 0.305 | - |
| SHV-75 | 1 (0.3) | 0 (0) | 1 (0.36) | - | - |
| SHV-76^ | 1 (0.3) | 0 (0) | 1 (0.36) | - | - |

Abbreviations: **aOR** (adjusted odds ratio), **ARGs** (Antimicrobial resistant genes), **CI** (confidence intervals), **OR** (odds ratio), **pCAI** (presumed community-acquired), **pHAI** (presumed hospital-associated).

ARGs: **CMY-4.v1** (CMY-4 beta-lactamase gene version 1), **DHA-1** (DHA-1 beta-lactamase), **CTX-M-15** (CTX-M-15 extended-spectrum beta-lactamase), **CTX-M-15*** (CTX-M-15 extended-spectrum beta-lactamase variant), **CTX-M-3** (CTX-M-3 extended-spectrum beta-lactamase), **NDM-1** (New Delhi metallo-beta-lactamase-1), **NDM-5** (New Delhi metallo-beta-lactamase-5), **OXA-1** (OXA-1 beta-lactamase), **OXA-1*** (OXA-1 beta-lactamase variant), **OXA-9.v1** (OXA-9 beta-lactamase version 1), **OXA-10** (OXA-10 beta-lactamase), **OXA-23** (OXA-23 beta-lactamase), **OXA-48** (OXA-48 beta-lactamase), **OXA-66** (OXA-66 beta-lactamase), **OXA-181** (OXA-181 beta-lactamase), **SHV-1** (SHV-1 beta-lactamase), **SHV-1^** (SHV-1 beta-lactamase variant), **SHV-11.v1^** (SHV-11 beta-lactamase version 1, variant), **SHV-11^** (SHV-11 beta-lactamase variant), **SHV-13*** (SHV-13 beta-lactamase variant), *SHV-172?** (SHV-172 beta-lactamase variant, possibly uncertain), **SHV-187** (SHV-187 beta-lactamase), **SHV-187*** (SHV-187 beta-lactamase variant), **SHV-187^** (SHV-187 beta-lactamase variant), **SHV-207** (SHV-207 beta-lactamase), **SHV-209*** (SHV-209 beta-lactamase variant), **SHV-220** (SHV-220 beta-lactamase), **SHV-26** (SHV-26 beta-lactamase), **SHV-27** (SHV-27 beta-lactamase), **SHV-28^** (SHV-28 beta-lactamase variant), **SHV-32** (SHV-32 beta-lactamase), **SHV-33** (SHV-33 beta-lactamase), **SHV-33*** (SHV-33 beta-lactamase variant), **SHV-36** (SHV-36 beta-lactamase), **SHV-61** (SHV-61 beta-lactamase), **SHV-62** (SHV-62 beta-lactamase), **SHV-75** (SHV-75 beta-lactamase), **SHV-76^** (SHV-76 beta-lactamase variant), **TEM-1D.v1** (TEM-1D beta-lactamase version 1), **TEM-33** (TEM-33 beta-lactamase), **TEM-34** (TEM-34 beta-lactamase), **VIM-1** (Verona integron-encoded metallo-beta-lactamase-1), and **SCO-1** (SCO-1 beta-lactamase).

pCAI isolates were defined as invasive KPn detected on admission or within 72 hours of hospitalization or if the death occurred in the community.

pHAI was defined as an invasive KPn detected more than 72 hours after admission to the hospital or if the DeCoDe panel attributed nosocomial infection to the causal pathway of the death.

Adjusted odd ratio and 95% CI calculated using logistic regression analyses. P-values of <0.05 are considered significant.

- too few variables to calculate

#### SUPPLEMENTARY FIGURES

##### a) Across all sites: Clonotypes

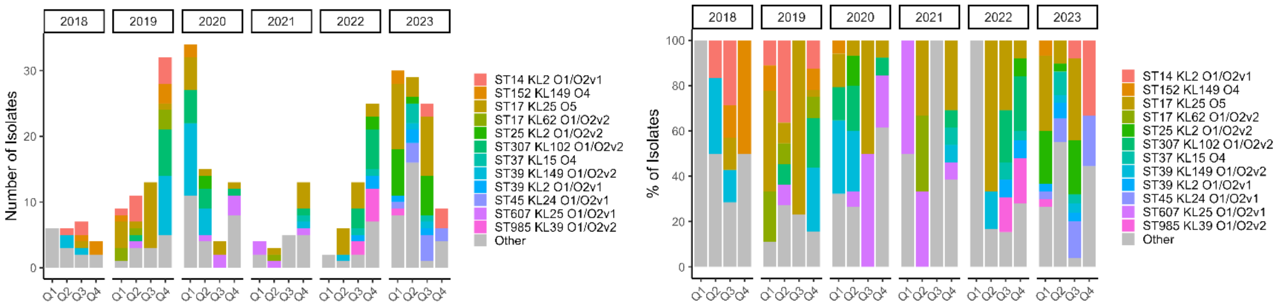


##### Across all sites: Sequence types

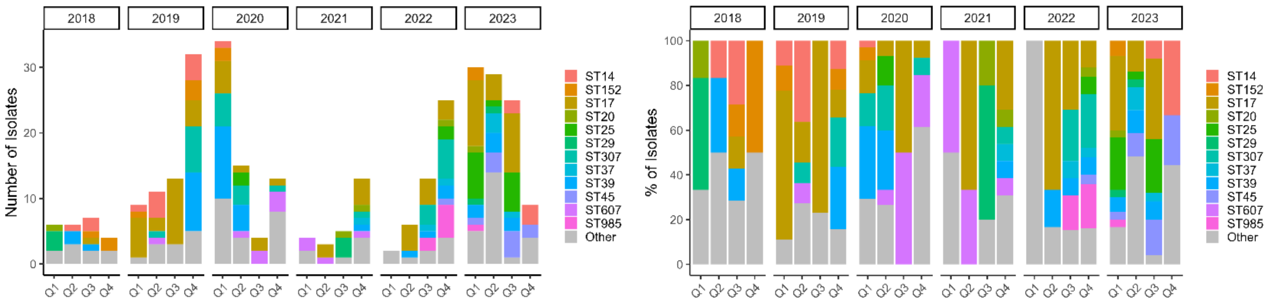


##### Across all sites: K-loci

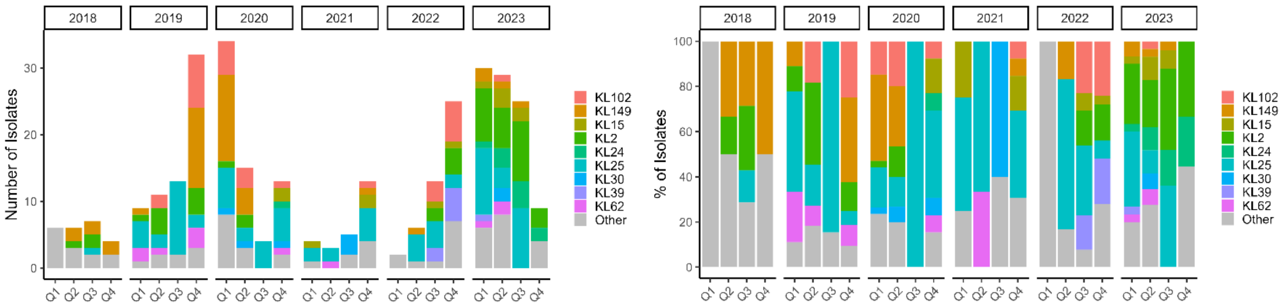


##### Across all sites: O-antigen types

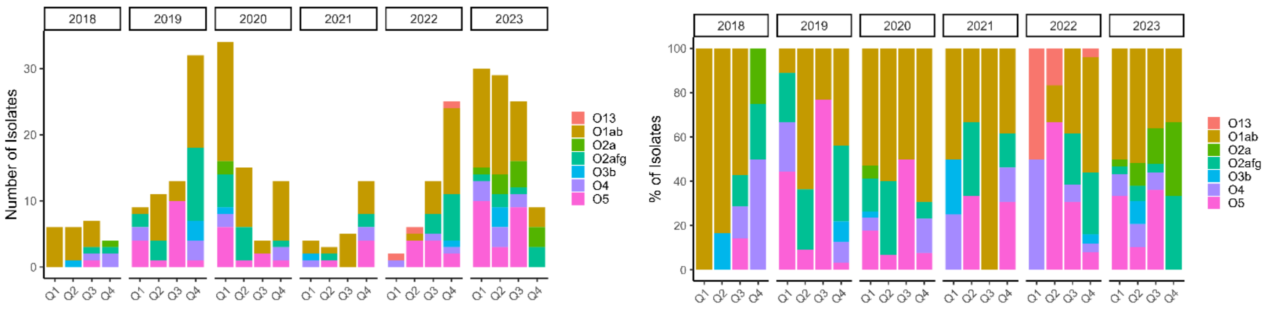

SUPPLEMENTARY

b) CHBAH: Clonotypes

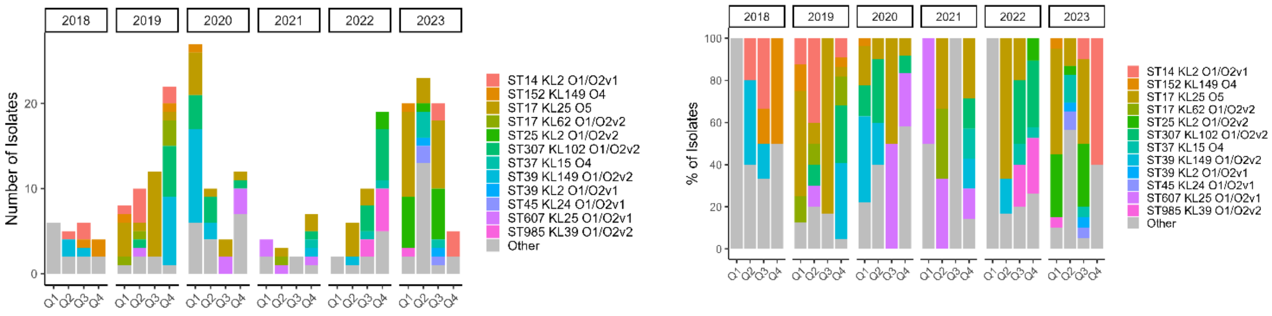


CHBAH: Sequence types

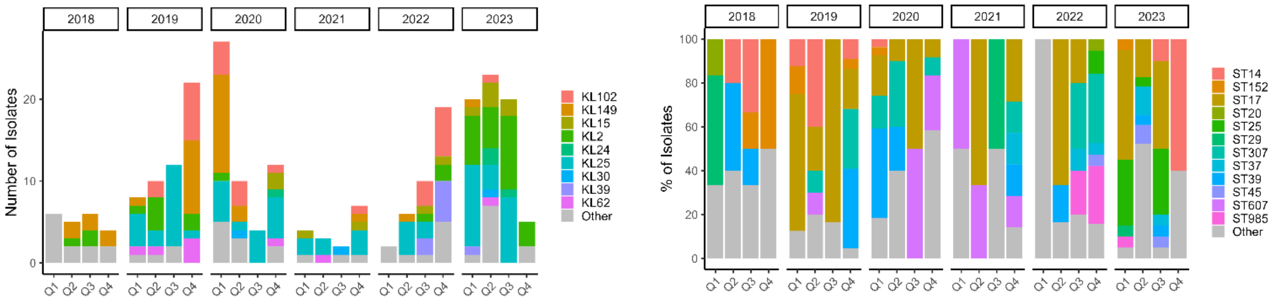


CHBAH: K-loci

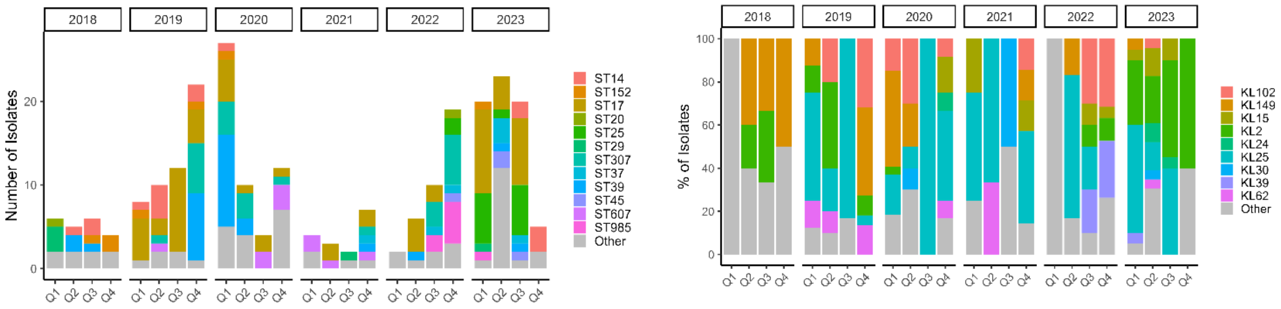


CHBAH: O-antigen types

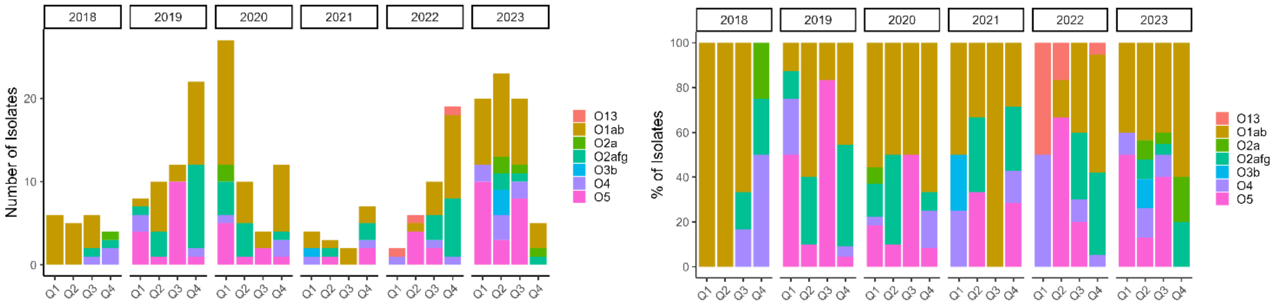

c) Leratong Hospital: Clonotypes

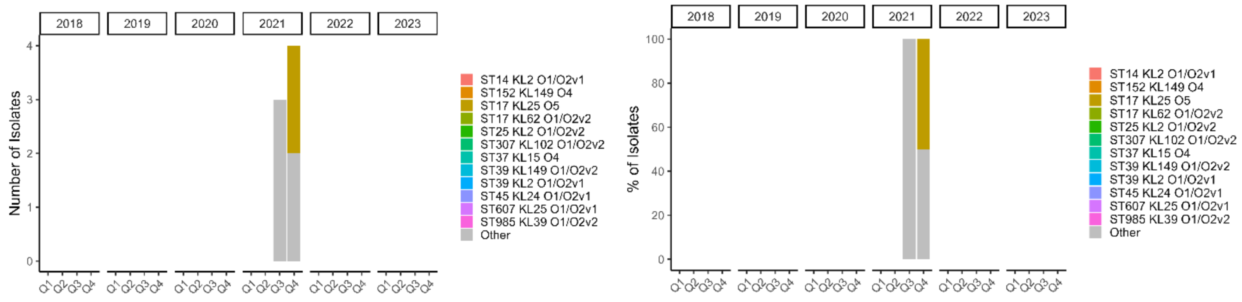


Leratong Hospital: Sequence types

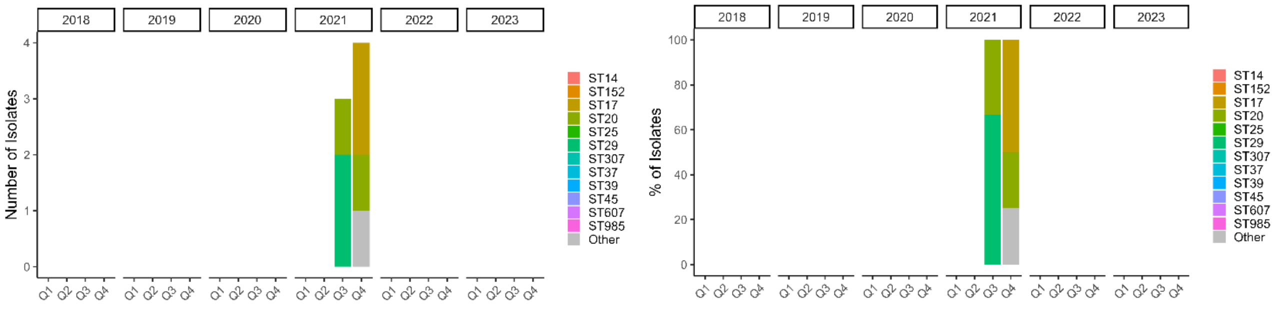


Leratong Hospital: K-loci

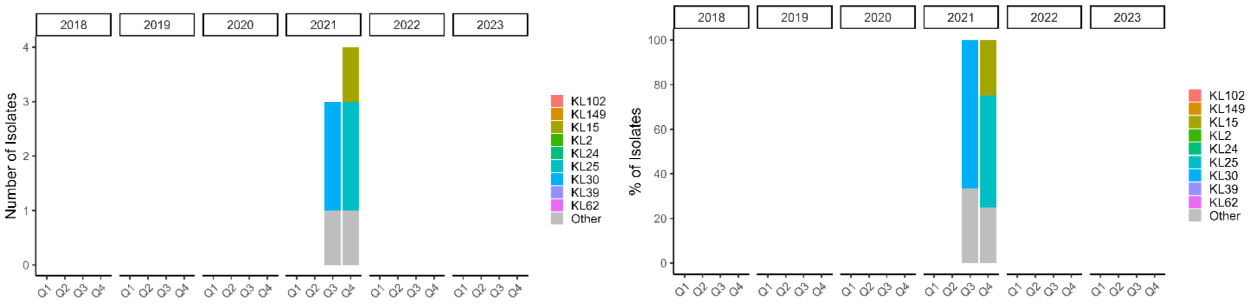


Leratong Hospital: O-antigen types

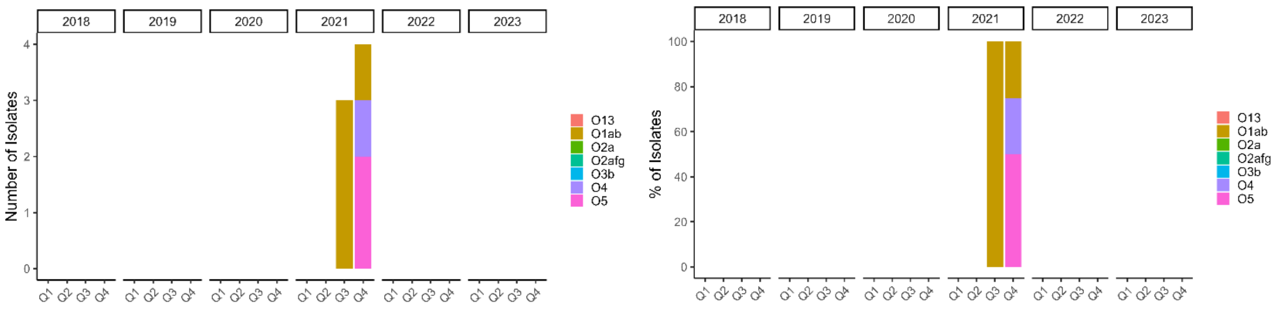

SUPPLEMENTARY

d) Rahima Moosa Mother and Child Hospital: Clonotypes

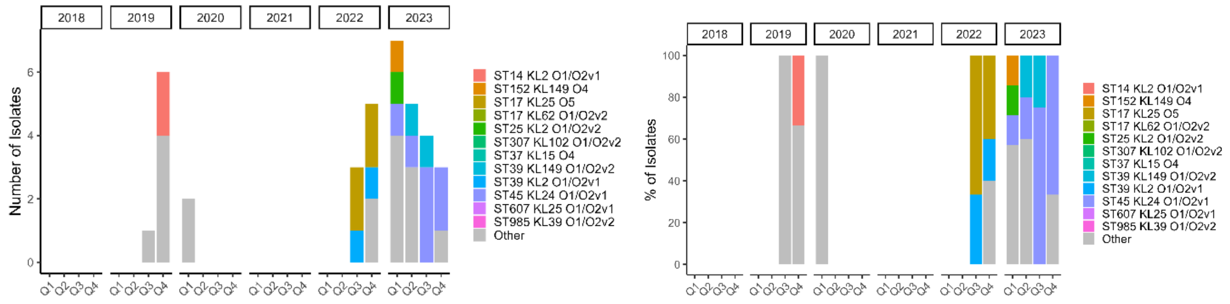


Rahima Moosa Mother and Child Hospital: Sequence types

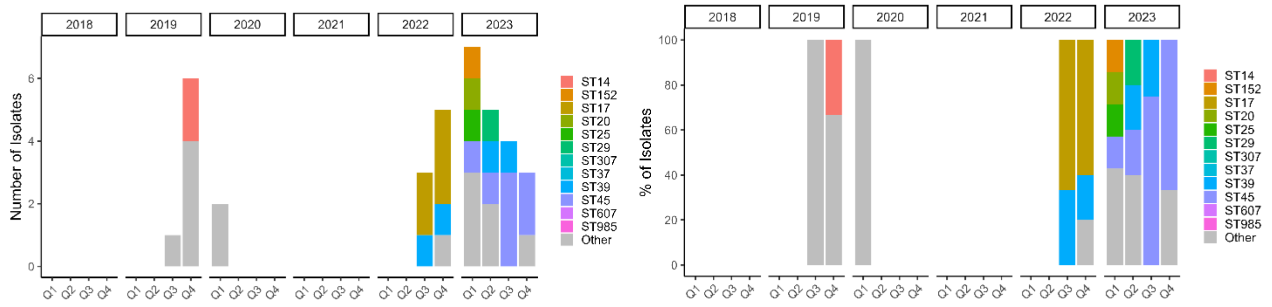


Rahima Moosa Mother and Child Hospital: K-loci

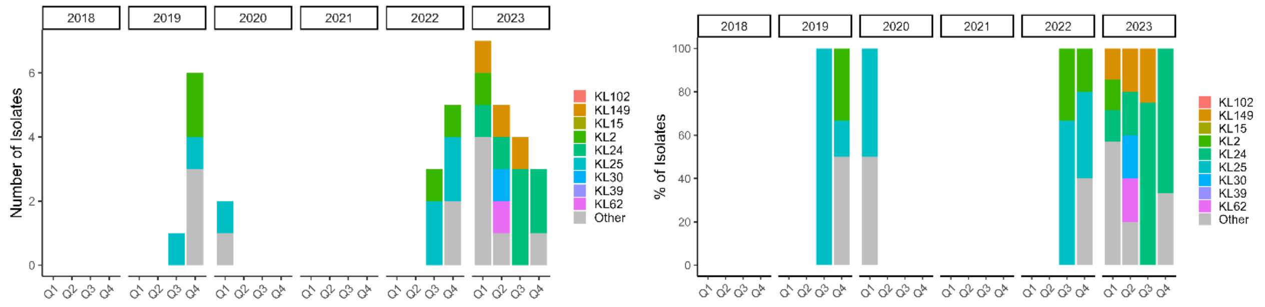


Rahima Moosa Mother and Child Hospital: O-antigen types

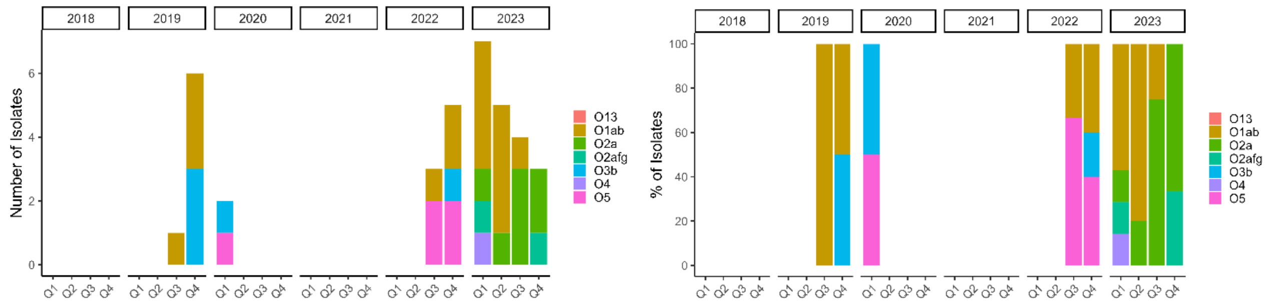

e) Steve Biko Academic Hospital: Clonotypes

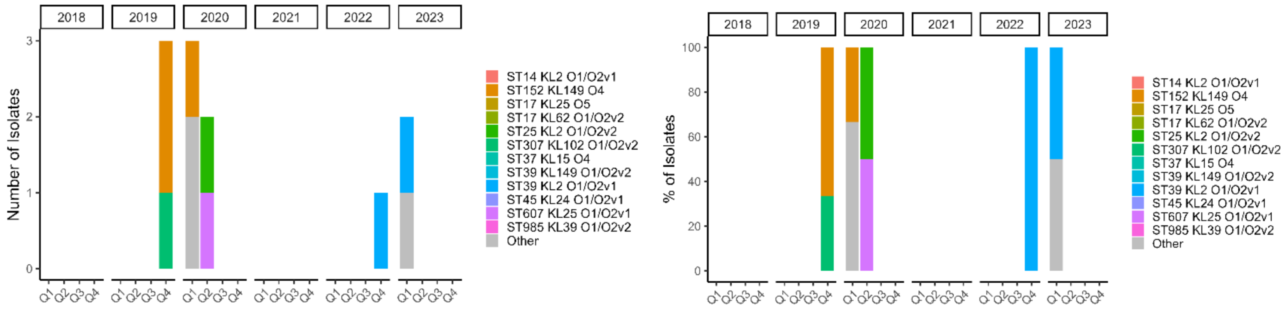


Steve Biko Academic Hospital: Sequence types

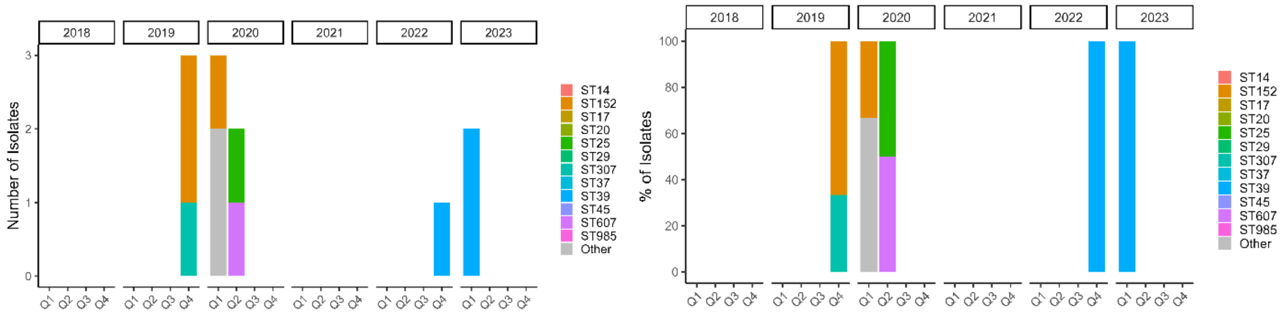


Steve Biko Academic Hospital: K-loci

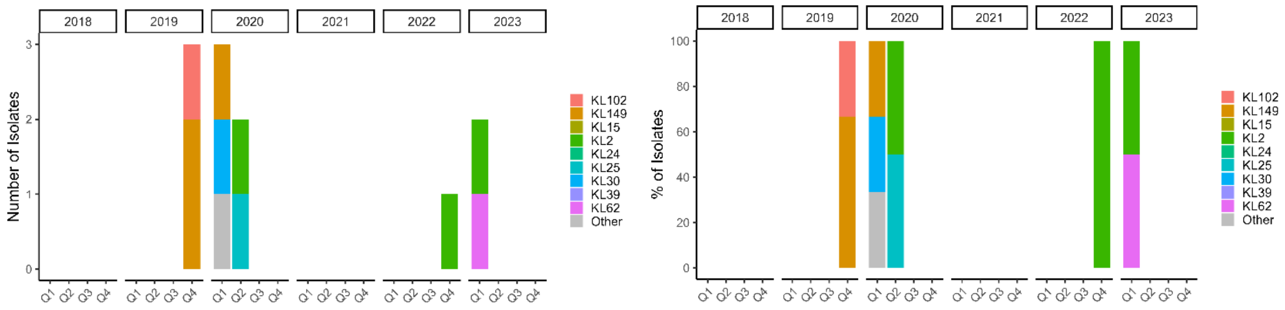


Steve Biko Academic Hospital: O-antigen types

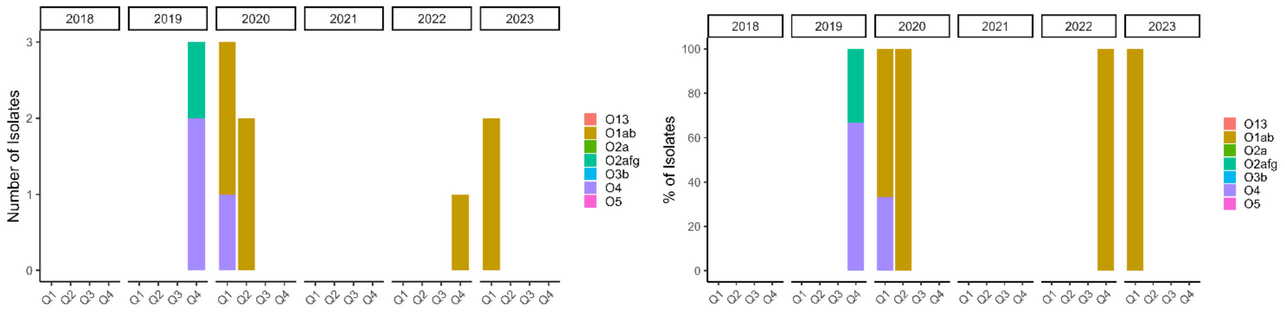


**Supplementary Figure 1:** Changes in the genotypes, ST, K-loci, and O-antigens of *K. pneumoniae* causing invasive disease in South African infants (a) across all sites, (b) Chris Hani Baragwanath Academic Hospital, (c) Leratong hospital, (d) Rahima Moosa Mother and Child Hospital, and (e) Steve Biko Academic Hospital over the study period.

**Alt text:** graphs showing the changes in the genotypes, sequence types (ST), K-loci, and O-antigens of *Klebsiella pneumoniae* causing invasive disease in South African infants. The data is presented for (a) all sites combined, (b) Chris Hani Baragwanath Hospital, (c) Leratong hospital, (d) Rahima Moosa Mother and Child Hospital, and (e) Steve Biko Academic Hospital between 2018 and 2023.

A.

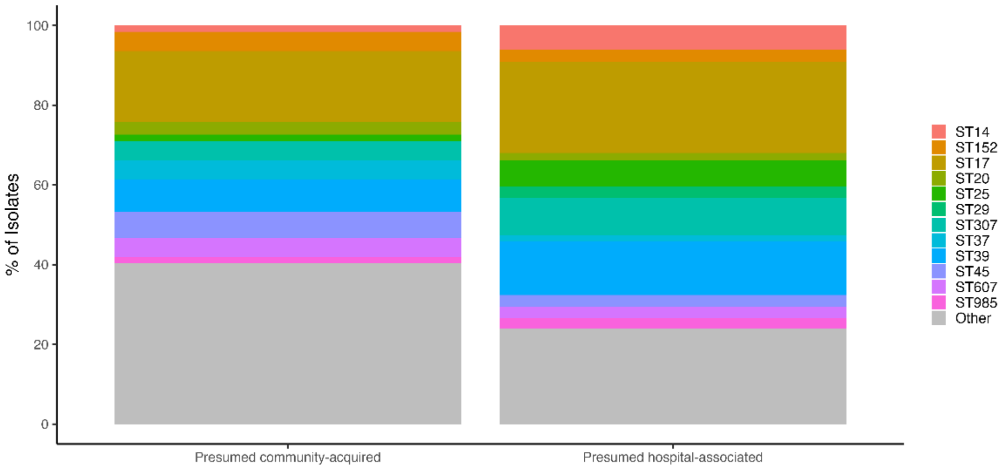


B.

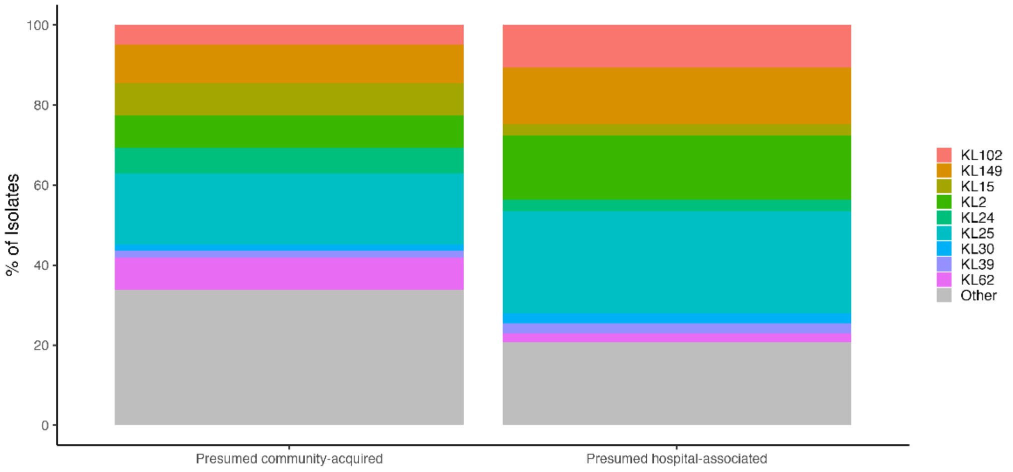


C.

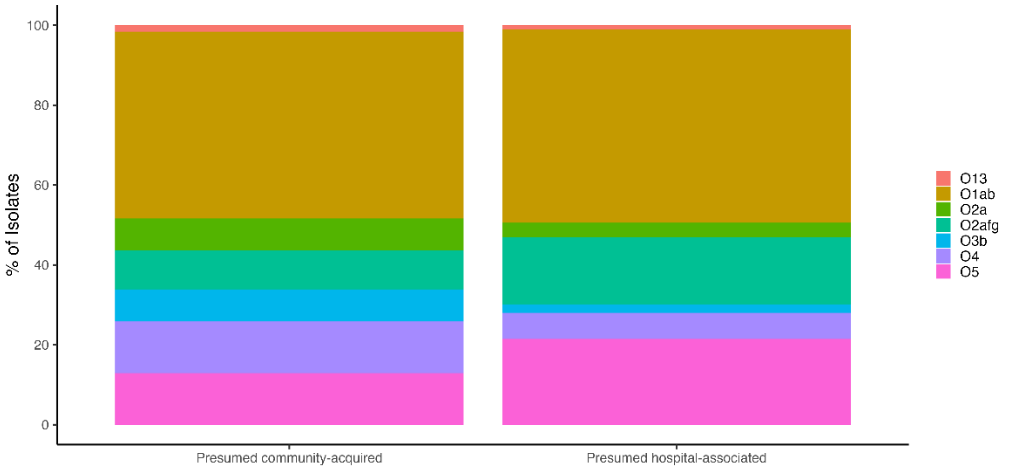


**Supplementary Figure 2:** Sequence types (a), K-loci (b), and O-antigens (c), by setting

**Alt text:** Graphs sowing the distribution of (a), sequence types (ST), (b) K-loci, and (c) O-antigens of *Klebsiella pneumoniae* causing invasive disease in South African infants. The data is stratified by cases presumed to be acquired in the hospital versus those assumed to be acquired in the community.

SUPPLEMENTARY

A.

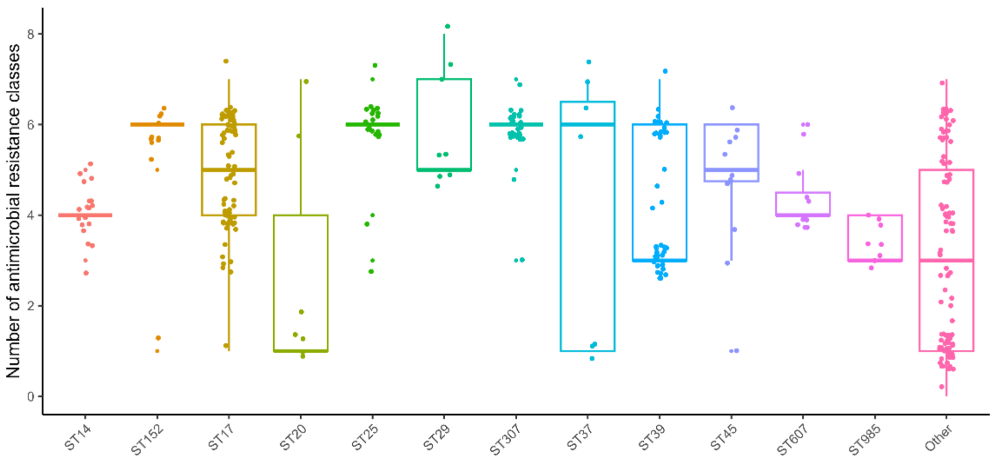


B.

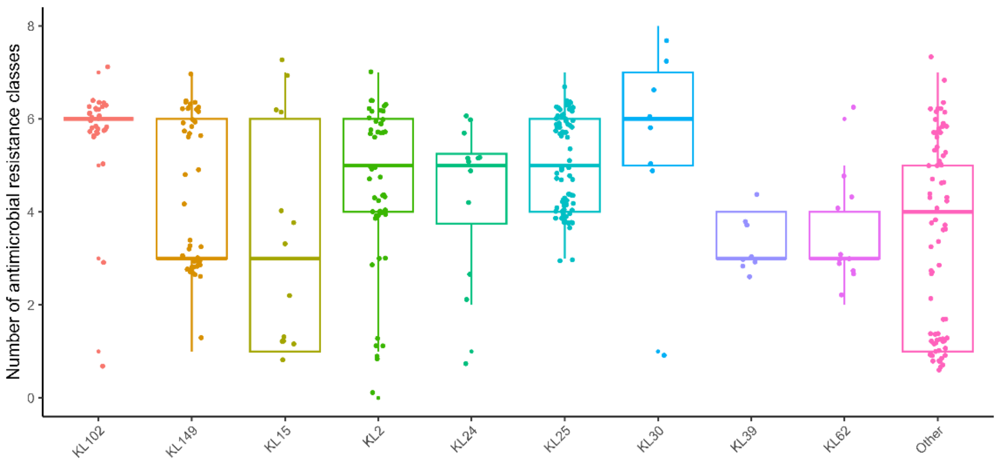


C.

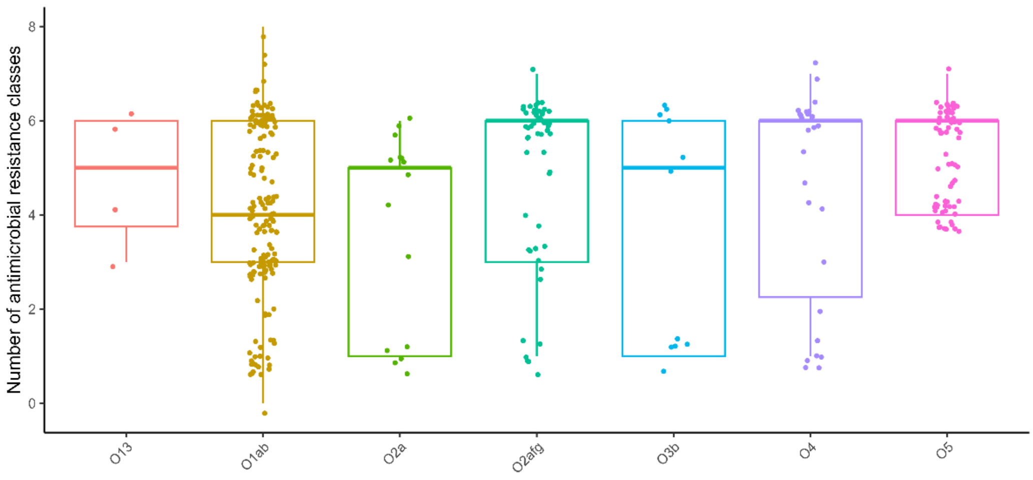

**Supplementary Figure 3:** Number of antimicrobial resistance classes, stratified by Sequence types (a), K-loci (b), and O-antigens (c).

**Alt text**: Graphs illustrating the number of different antimicrobial classes to which the *Klebsiella pneumoniae* strains were resistant. The data is stratified by the most common (a) Sequence types, (b) K-loci, and (c) O-antigens.
